# Modelling lockdown-induced 2^nd^ COVID waves in France

**DOI:** 10.1101/2020.06.24.20139444

**Authors:** Jean Daunizeau, Rosalyn Moran, Jules Brochard, Jérémie Mattout, Richard Frackowiak, Karl Friston

## Abstract

As with the Spanish Flu a century ago, authorities have responded to the current COVID-19 pandemic with extraordinary public health measures. In particular, lockdown and related social distancing policies are motivated in some countries by the need to slow virus propagation—so that the primary wave of patients suffering from severe forms of COVID infection do not exceed the capacity of intensive care units. But unlocking poses a critical issue because relaxing social distancing may, in principle, generate secondary waves. Ironically however, the dynamic repertoire of established epidemiological models that support this kind of reasoning is limited to single epidemic outbreaks. In turn, predictions regarding secondary waves are tautologically derived from imposing assumptions about changes in the so-called “effective reproduction number”. In this work, we depart from this approach and extend the LIST (Location-Infection-Symptom-Testing) model of the COVID pandemic with realistic nonlinear feedback mechanisms that under certain conditions, cause lockdown-induced secondary outbreaks. The original LIST model captures adaptive social distancing, *i*.*e*. the transient reduction of the number of person-to-person contacts (and hence the rate of virus transmission), as a societal response to salient public health risks. Here, we consider the possibility that such pruning of socio-geographical networks may also temporarily isolate subsets of local populations from the virus. Crucially however, such unreachable people will become susceptible again when adaptive social distancing relaxes and the density of contacts within socio-geographical networks increases again. Taken together, adaptive social distancing and network *unreachability* thus close a nonlinear feedback loop that endows the LIST model with a mechanism that can generate autonomous (lockdown-induced) secondary waves. However, whether and how secondary waves arise depend upon the interaction with other nonlinear mechanisms that capture other forms of transmission heterogeneity. We apply the ensuing LIST model to numerical simulations and exhaustive analyses of regional French epidemiological data. We find evidence for this kind of nonlinear feedback mechanism in the empirical dynamics of the pandemic in France. However, rather than generating catastrophic secondary outbreaks (as is typically assumed), the model predicts that the impact of lockdown-induced variations in population susceptibility and transmission may eventually reduce to a steady-state endemic equilibrium with a low but stable infection rate. In brief, except if immunity is lost (because of, e.g., virus mutations), a secondary COVID wave before winter is unlikely.

## 1. Introduction

At the time of writing, most countries in the world have engaged in stringent public health measures, including lockdown, to combat the COVID pandemic outbreak. Lockdown is motivated by a need to slow virus propagation such that the first wave of patients—suffering from severe forms of COVID—does not exceed the capacity of intensive care units. In the absence of pharmacological treatment, and given the paucity of scientific knowledge about the biomedical features of this pandemic, such a strategy was very reasonable (Lau *et al*., 2020; Roux *et al*., 2020; Thornton, 2020). Now, given the devastating economic cost of lockdown (Inoue and Todo, 2020), exit strategies are being discussed and implemented (Peto *et al*., 2020). The issue here is that relaxing social distancing may, in principle, generate secondary outbreak waves (Domenico *et al*., 2020a; Lawton, 2020). Evidence for such a scenario was observed a century ago, in some US cities that attempted to lift lockdown at critical times during the Spanish Flu pandemic (Johnson and Mueller, 2002). In fact, oscillations in influenza infection rates were retrospectively shown to two have two distinct causes: (i) short-term, lockdown-induced, changes in virus susceptibility and transmission rates, and (ii) longer-term losses of immunity that originated from virus mutations (Martini *et al*., 2019; Phillips, 2014; Reid *et al*., 2001). Crucially however, only the latter was shown to cause catastrophic secondary outbreaks at the national level (Moxnes and Christophersen, 2008; Spinney, 2018). This raises the question of whether and how lockdown can induce secondary COVID outbreak waves.

In the context of the COVID pandemic, most established epidemiologic models that are used to support this kind of prediction do not consider opponent nonlinear feedback mechanisms that might contribute to the generation and/or attenuation of such secondary waves. In most instances, current model-based predictions work by imposing assumptions about abrupt increases in the so-called “effective reproduction number” (**R**) that might follow from relaxing lockdown and/or other related social distancing measures (Balabdaoui and Mohr, 2020; Domenico *et al*., 2020b; Flaxman, 2020; Hazem *et al*., 2020; Leung *et al*., 2020; Moghadas *et al*., 2020; Sarma and Ghosh, 2020; Xu and Li, 2020, 2020). From a dynamical systems theoretical perspective, this process is tautological; the aim should be to predict oscillations of **R** by an exhaustive consideration of basic, possibly opponent, socio-epidemiological mechanisms. This limitation is a feature of models that are simple extensions of the SIR model (Kermack *et al*., 1927), whose dynamical repertoire is limited to single epidemic outbreaks, once their parameters are set (Eubank *et al*., 2020).

We depart from this approach to extend a recent dynamic causal model of the COVID pandemic (Friston *et al*., 2020a). In brief, the LIST (Location-Infection-Symptom-Testing) model captures many nonlinear mechanisms that are *a priori* relevant for predicting non-trivial features of population responses to the COVID virus. These include, but are not limited to, heterogeneities in susceptibility and transmission, whose impact explain between-country differences in the dynamics of the primary COVID outbreak (Friston *et al*., 2020b, 2020c, 2020d). It also provides a quantitative description of adaptive social distancing, *i*.*e*., the transient reduction of person-to-person contacts (and hence, virus transmission speed) that results from societal attitudes towards salient public health risks. This makes the rise and fall of social distancing an internal process in the model, which can be used to assess the economic cost of the COVID pandemic (in terms of, *e*.*g*., lost working days). In addition, we consider the possibility that such lockdown-induced pruning of socio-geographical networks may also temporarily isolate subsets of the population from the virus. Crucially, such “unreachable*”* people will become susceptible again when adaptive social distancing eventually relaxes. Taken together, adaptive social distancing and network *unreachability* thus close a nonlinear feedback loop that endows the LIST model with a mechanism that can generate autonomous (lockdown-induced) secondary waves. However, whether and how lockdown-induced secondary waves arise depends upon the balance between this and other mitigating mechanisms included in the LIST model.

In what follows, we first summarize the basics of SIR modelling and compare it to LIST dynamics. We then show how to extend the LIST model with *network unreachability*. Finally, we demonstrate the inferences and predictions that can be made from quantifying the relative contribution of LIST-like opponent nonlinear feedback mechanisms from available epidemiological data. We report results from an exhaustive LIST analysis of French regional data and conclude with predictions regarding infection and acquired immunity levels in France in the future.

## 2. Standard epidemiological models

### a. The basic SIR framework

Standard epidemiologic models are variants of the basic SIR framework (Kermack *et al*., 1927). In brief, these models assume that the population is divided into, ‘Susceptible’, ‘Infected’, and ‘Removed^1^’ compartments, through which individuals transit at a pace that is characteristic of the time course of the infection modified by socio-medical measures (see Figure 1 below).

**Figure 1:**
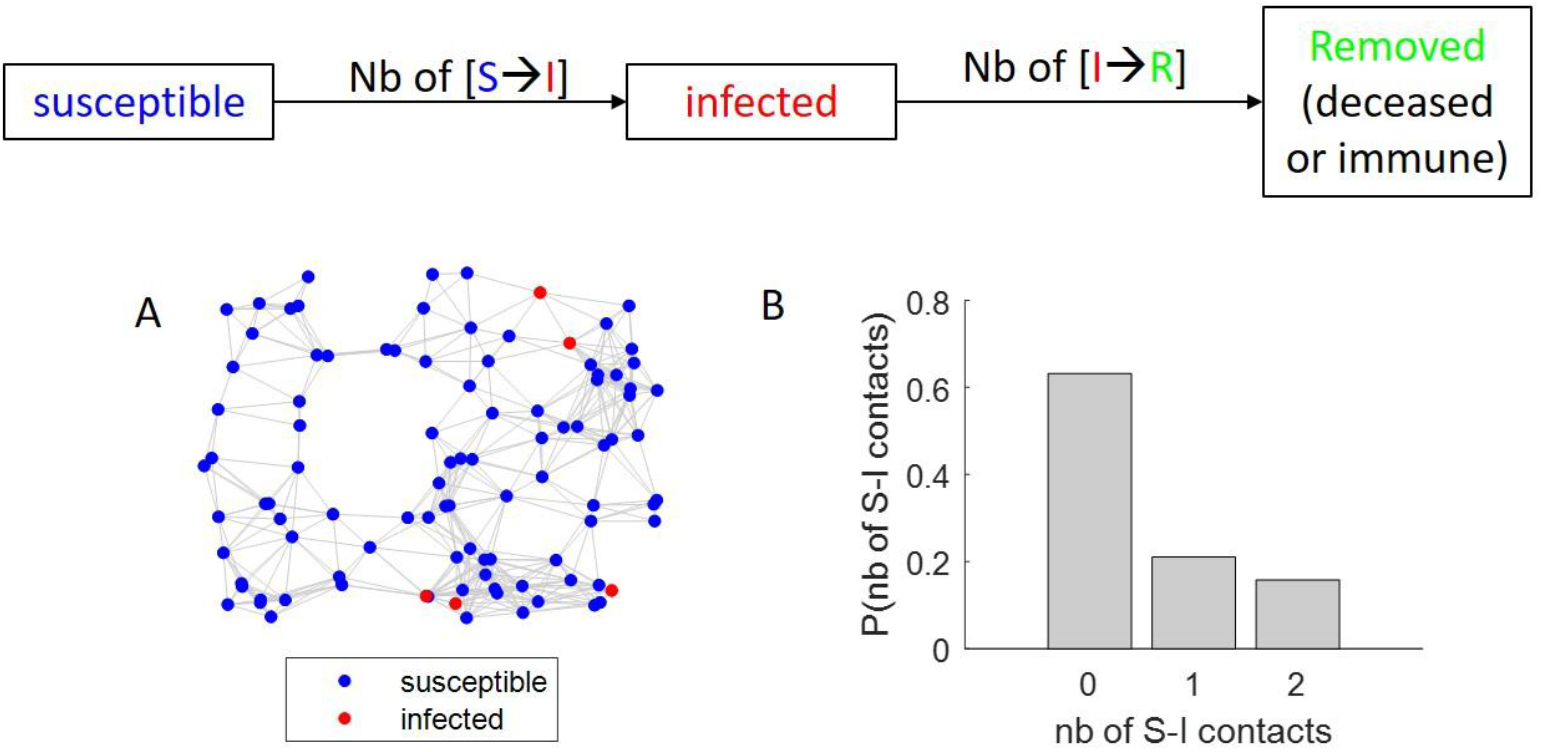
The basic SIR model. A: Example of a socio-geographical network relating people with each other (red dots: infected people, blue dots: susceptible people). B: Probability, for susceptible individuals in the toy socio-geographical network of panel A, of being in direct contact with someone who is infected.

Let *N, S, I*, and *R* be the total population size, the number of susceptible people, the number of infected people and the number of removed (either immune or dead) people, respectively. The main contribution of SIR models is to recycle basic statistical physics derivations to predict the number of people who transit from the susceptible compartment to the infected compartment.

Let *P* (*S* → *I*) be the probability that one susceptible individual will become infected:

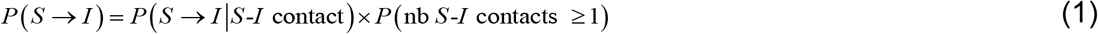

Where *P* (*S* → *I S* -*I* contact) and *P* (nb *S* -*I* contacts ≥ 1) are the probabilities that someone susceptible will become infected if (s)he is in contact with an infected person (disease infectiousness) and that someone susceptible is in direct contact with at least one infected person, respectively. Using a simple mean-field approach, the latter can be written as:

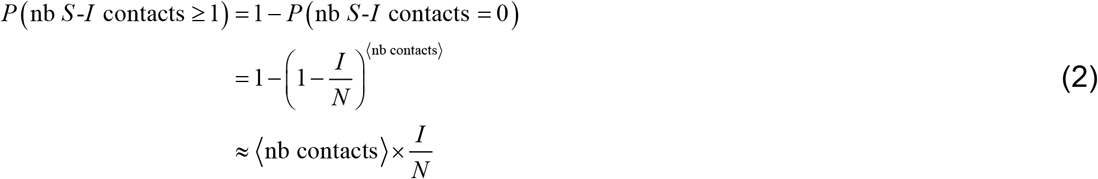

where ⟨ nb contacts ⟩ is the average number of direct contacts between individuals in the population. Inserting Equation 2 into Equation 1 yields:

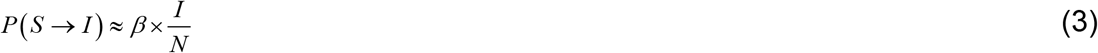

where

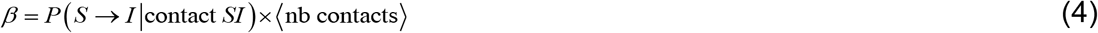

is an unknown parameter of the model. The number of people who become infected is then simply given by *S* × *P* (*S* → *I*).

To complete the SIR model, one then simply expresses the number of infected people who are ‘removed’ as *γ* × *I*, where *γ* = *P* (*I* → *R*) is the (unknown) probability that one infected person will be removed.

This then enables one to write the following balanced system of ordinary differential equations:

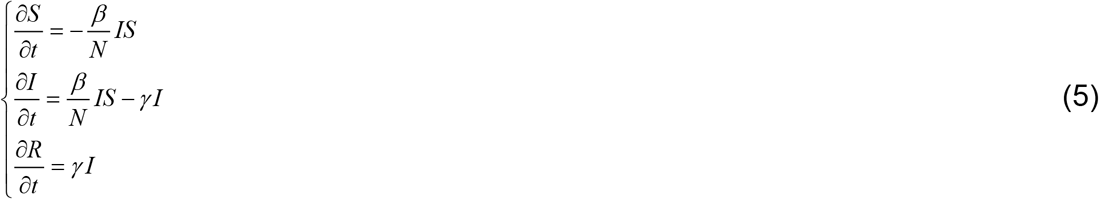

Note that *β* and *γ* can be thought of as rate constants. For example, *γ* effectively controls the time it takes for one infected individual to be removed (more formally, it is the inverse half-life of exponential I→ S decay dynamics).

Figure 2 below shows an exemplary simulated wave of an epidemic as people become infected and subsequently immune or dead.

**Figure 2:**
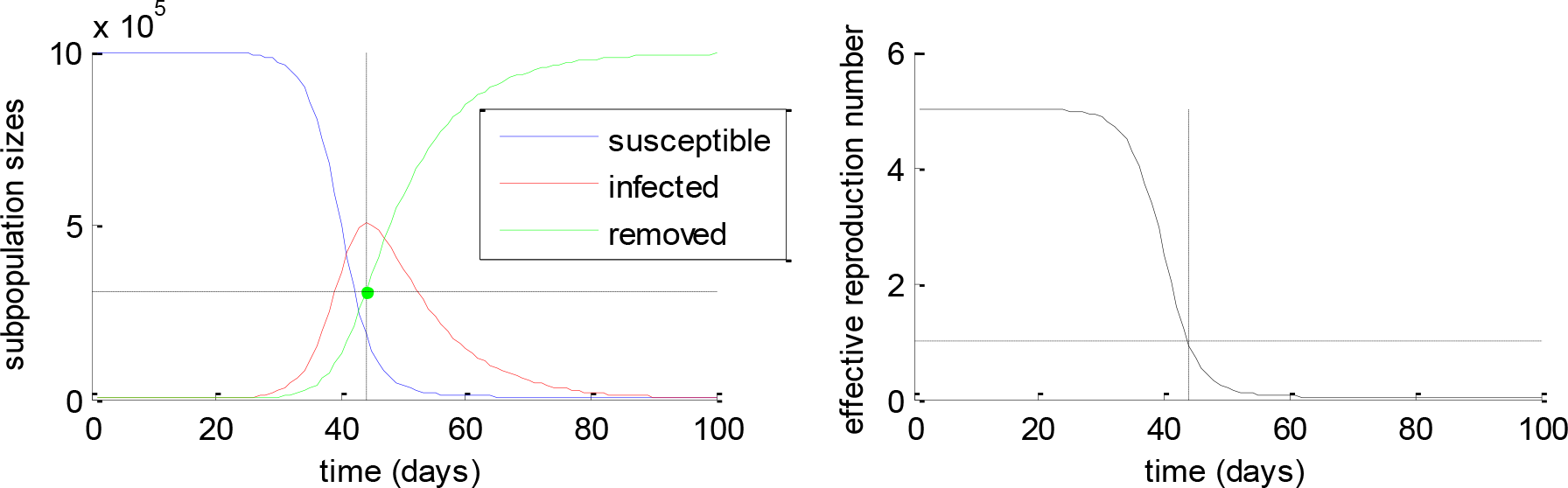
Simulated SIR epidemic wave. Left: SIR compartment sizes (y-axis, blue: susceptible, red: infected, green: removed, total population size: N=10^6^) are plotted as a function of time (x-axis). Right: The effective reproduction number (y-axis) is plotted as a function of time (x-axis).

Now recall that the effective reproduction number **R** quantifies the expected number of people who will be directly infected by a typical contagious person. In the context of SIR models, one can show that **R** reduces to a simple function of the SIR parameters, *i*.*e*. **R≜***β S γ N* (Eubank *et al*., 2020). When **R** becomes smaller than one, then the number of new infected people decreases, which signals that the epidemic wave has reached a peak. The proportion of immune people when this happens is called “herd immunity*”*, which effectively is the immunity level that needs to be attained for the epidemic to stop growing. In the simulated example above, herd immunity is achieved at about one third of the total population size.

The critical issue here is that SIR models invariably predict that the population dynamics will show a single transient increase in infection rate (epidemic outbreak), and eventually reach a so-called “disease-free equilibrium” (exhaustion of the ‘susceptible’ compartment)^2^. In other words, the dynamical repertoire of SIR models is limited to single outbreaks. Under the SIR model, lockdown-induced secondary waves thus arise from exogenous changes in model parameters that are set to co-occur with lockdown onsets and offsets (Balabdaoui and Mohr, 2020; Domenico *et al*., 2020b; Hazem *et al*., 2020; Leung *et al*., 2020; Moghadas *et al*., 2020; Sarma and Ghosh, 2020; Xu and Li, 2020, 2020). Typically, efficient lockdown is viewed as a significant reduction in the number of person-to-person contacts. This pruning of socio-geographical networks is typically modelled by reducing *β* (cf. Equation 4) to induce arbitary small effective reproduction numbers (**R** < 1). In turn, this yields an unstable steady-state whereby the infection rate is maintained at a low level (*I* ≈ 0 equilibrium) as long as lockdown is effective. Under this kind of theoretical scenario, a post-lockdown secondary wave is inevitable if lockdown measures are lifted before herd immunity has been reached (and *β* returns to its pre-lockdown value). This is why early epidemiological COVID reports recommended lockdown for as long as 16 months (Flaxman, 2020).

### b. Extended SIR models

Many extensions to the 100-year old SIR model have been proposed since. In brief, most of them split the ‘removed’ compartment into ‘immune’ and ‘deceased’ sub-states. They also consider biomedical specificities of the disease (*e*.*g*., in terms of medical care). In addition, they typically stratify the population into age and gender groups, because these have different characteristics in terms of number of person-to-person contacts, susceptibility to infection and prognostic risk. Although these kinds of extensions significantly improve the realism of SIR models, they still do not capture feedback mechanisms that can generate and/or attenuate secondary waves. In brief, most extended SIR models continue to work in a *feedforward* manner, whereby people transit from one model compartment to the next, until the initial pool of susceptible people is exhausted. As an example, Figure 3 shows the structure of a very recent SIR model that was applied to the current French component of the COVID pandemic (Salje *et al*., 2020).

**Figure 3:**
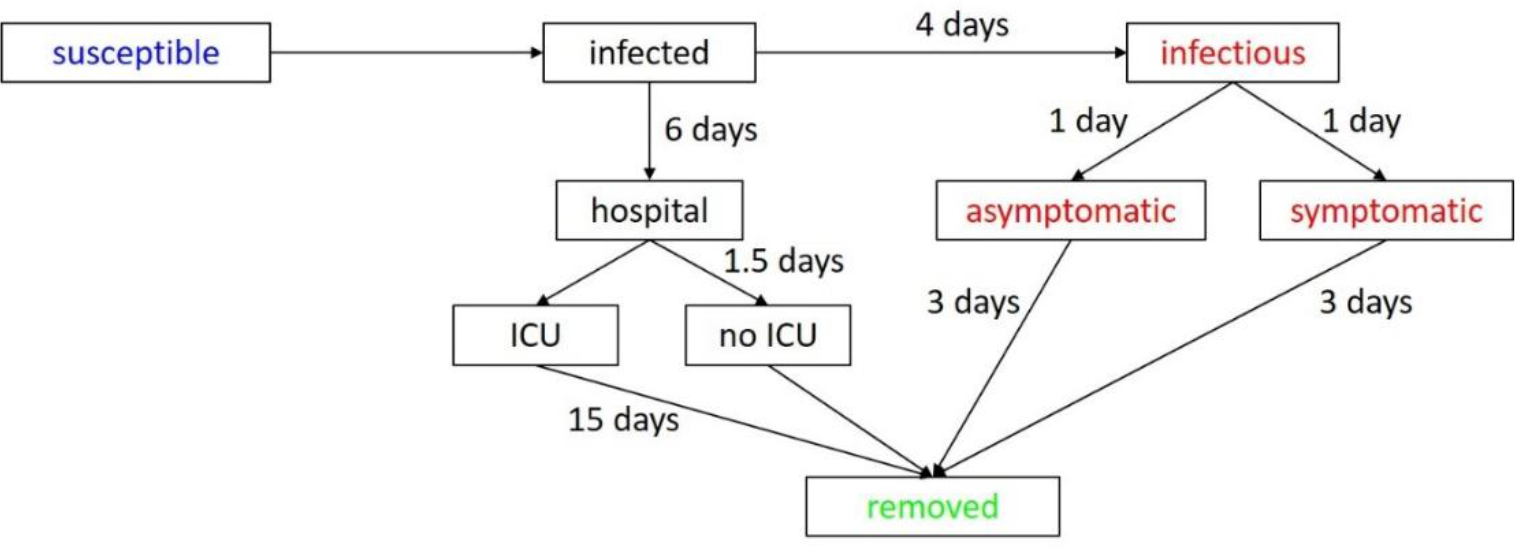
Recent extended SIR model structure. The model includes a set of ‘pre-infectious’, symptomatic/asymptomatic and hospital care sub-states with transition dynamics controlled by rate constants that are set according to COVID-specific biomedical data. Adapted from the supplementary material of (Salje *et al*., 2020).

Figure 4 shows the results of French data analysis based upon this extended SIR model (Salje *et al*., 2020) Crucially, this model predicts that as of the 11^th^ of May the proportion of infected people in the French population is between 2% and 10% (cf. panel F). Hence, the authors conclude that a post-lockdown secondary outbreak is highly likely because herd immunity has not been reached, *i*.*e*. most people remain susceptible to the virus. At the time of writing, this secondary wave has not happened. Under such extended SIR models, the only possible explanation is that the combination of efficient individual and institutional countermeasures (mask-wearing, limited public gatherings, “test and trace” processes, *etc*) still somehow maintain the unstable *I* ≈ 0 equilibrium. However, this seems at odds with the limited population adherence to self-constraining behaviour that has been seen since lockdown was lifted in France^3^.

**Figure 4:**
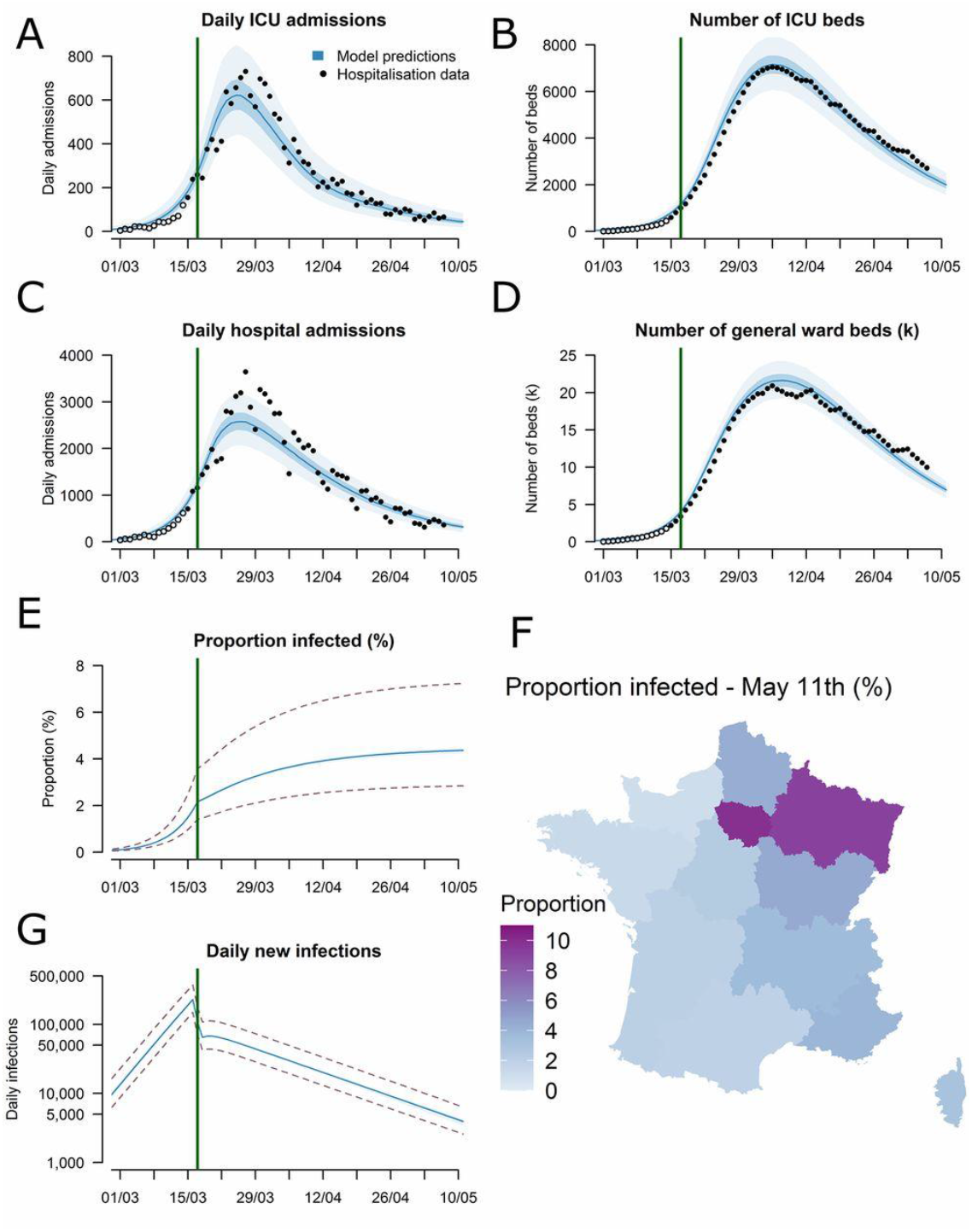
Results of the extended SIR model for France (Salje *et al*., 2020). A: Daily admissions to ICUs in metropolitan France. B: Number of ICU beds occupied. C: Daily hospital admissions. D: Number of general ward beds occupied. E: Predicted proportion of the population infected (the green vertical line indicates the lockdown onset). F: Predicted proportion of the population infected by 11 May 2020 for each of the 13 regions of metropolitan France. G: Daily new infections in metropolitan France (logarithmic scale).

As we will see below, the LIST model suggests a qualitatively different scenario for the relationship between lockdown and infection rates.

## 3. The LIST model

### a. A brief introduction to the factorial LIST model

In what follows, we briefly describe a novel generalisation of SIR models: namely, the LIST model (Friston *et al*., 2020a, 2020c, 2020d). In brief, the LIST model considers four interacting factors describing location, infection status, symptom/clinical status and test status respectively, where the infection factor can be thought of as an extended SIR model. Within each factor people transition probabilistically between (four to five) distinct states. Given a set of 24 model parameters (see below), the model describes the temporal dynamics of the marginal probabilities of belonging to any state within each factor. The location factor describes if an individual is at home, at work^4^, in an intensive care unit (ICU) or in the mortuary. Infection status is the closest factor to native SIR models, and includes susceptible, infected, contagious, resistant, or immune states. The distinction between the two latter sub-states will become clearer below. The clinical status factor comprises asymptomatic, symptomatic, acute respiratory distress syndrome (ARDS) or deceased states. Finally, the diagnostic status captures the fact that a given individual can be untested, waiting for the results of a test, or found to be either positive or negative. Model transitions amongst states are controlled by time constants (*e*.*g*., the contagious period) and probability constants (*e*.*g*., the probability of dying when in ICU). Critically, these transitions do not impose a feedforward structure to the model.

For example, people who acquire immunity may lose it and become susceptible again (though this may happen with a timescale of months or even years). The ensuing set of state probabilities can then be related to some specific observable epidemiologic outcomes, such as the number of deceased people per day or the number of people newly infected who have been tested positive. Figure 5 summarizes the causal structure implicit in conditional transition probabilities. We refer the reader to Friston *et al*. (Friston *et al*., 2020a) for a complete mathematical description of the model.

**Figure 5:**
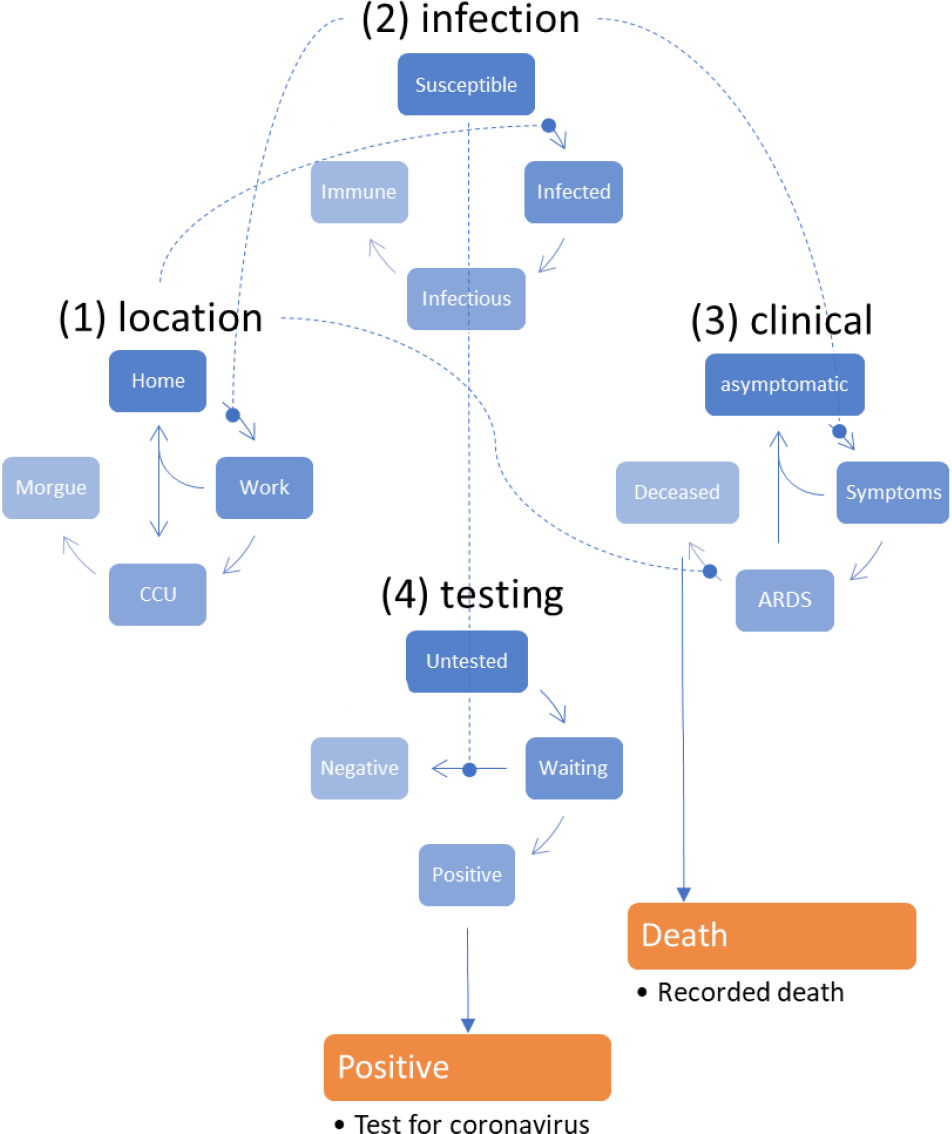
Causal structure of the original LIST model (adapted from Friston *et al*. 2020). In brief, the original LIST model assumes that (i) there is a progression from a state of susceptibility to immunity, through a period of (pre-contagious) infection to an infectious (contagious) status, (ii) there is a progression from asymptomatic to ARDS, where people with ARDS can either recover to an asymptomatic state or die, (iii) people can move from home to work or to ICU if they develop an ARDS, and (iv) the testing status progresses from untested, to waiting for results, to being declared positive or negative. With this setup, one can be in any location, with any infectious status, expressing symptoms or not and having test results or not. Note that—in this construction—it is possible to be infected and yet be asymptomatic. Crucially, the transitions within any factor depend upon the marginal distribution of other factors. For example, the probability of becoming infected, given that one is susceptible to infection, depends upon whether one is at home or at work (because this determines the number of person-to-person contacts). Similarly, the probability of developing symptoms depends upon whether one is infected or not. The probability of being tested negative depends upon whether one is susceptible (or immune) to infection, and so on. Finally, to complete the circular dependency, the probability of leaving home to go to work depends upon the number of symptomatic people in the population, mediated by social distancing. We will see that this feedback is critical for generating autonomous oscillatory modes of disease propagation. At any point in time, the probability of being in any combination of the four states determines what would be observed at the population level. For example, the occupancy of the ‘deceased’ level of the clinical factor determines the current number of recorded deaths. Similarly, the occupancy of the ‘positive’ level of the testing factor determines the reported number of positive cases. Note that this schematic does not include the ‘resistant’ state of the infection status factor (more on this later). A complete mathematical description of the LIST model can be found in the Appendix 1.

Parameter estimation and model comparison relies on a variational Bayesian scheme (Daunizeau, 2018; Friston *et al*., 2007) which is adopted in established computational neuroscience toolboxes (Ashburner, 2012). In addition to semi-informed prior distributions, model inversion—in the current implementation—places hard constraints on parameters to ensure that they stay within admissible ranges. More precisely, rate constants and probability constants are derived by passing unbounded parameters through exponential and sigmoid mappings, respectively^5^. Table 1 below recapitulates the unknown model parameters, in terms of their prior means and associated hard constraints.

**Table 1:**
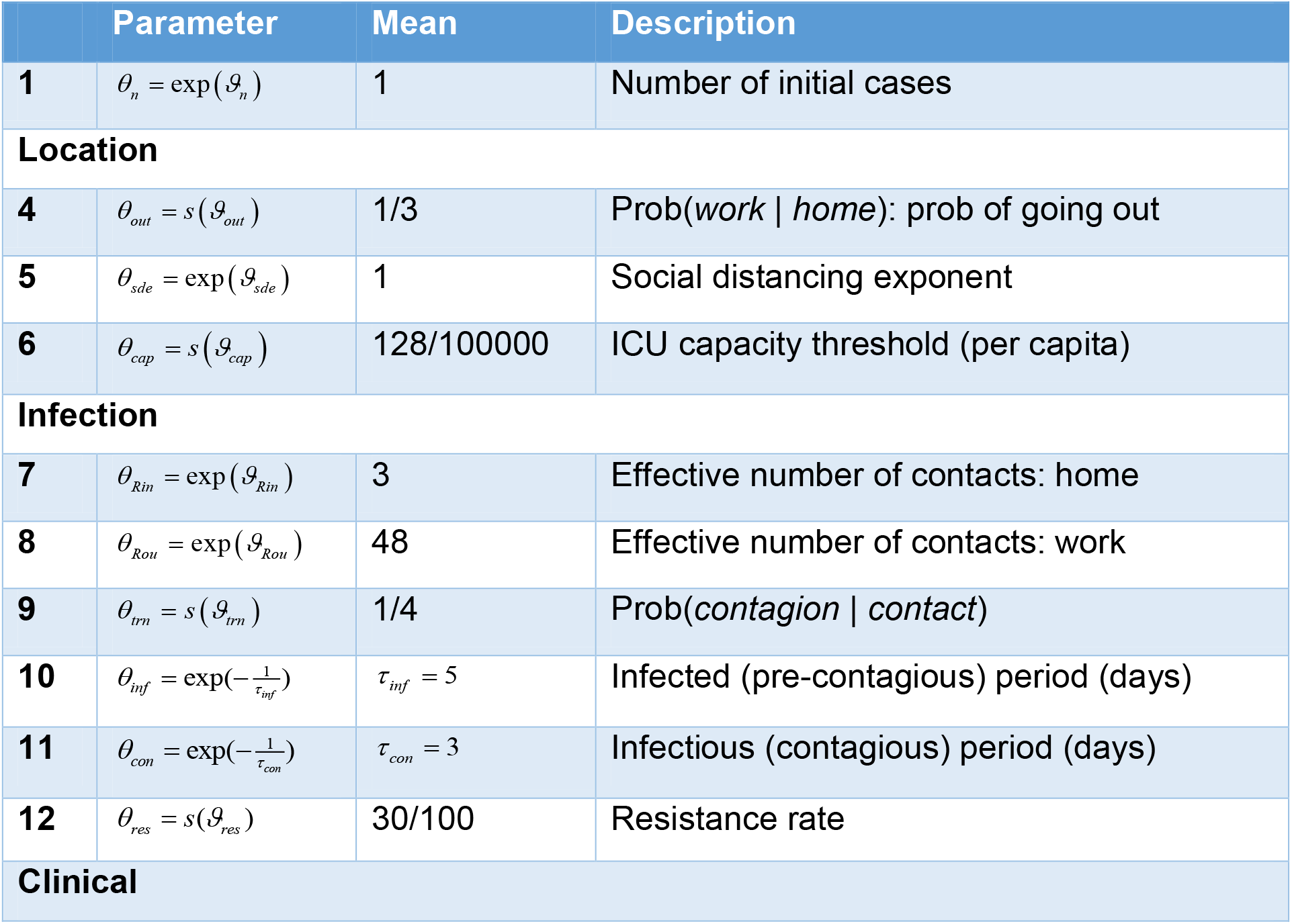
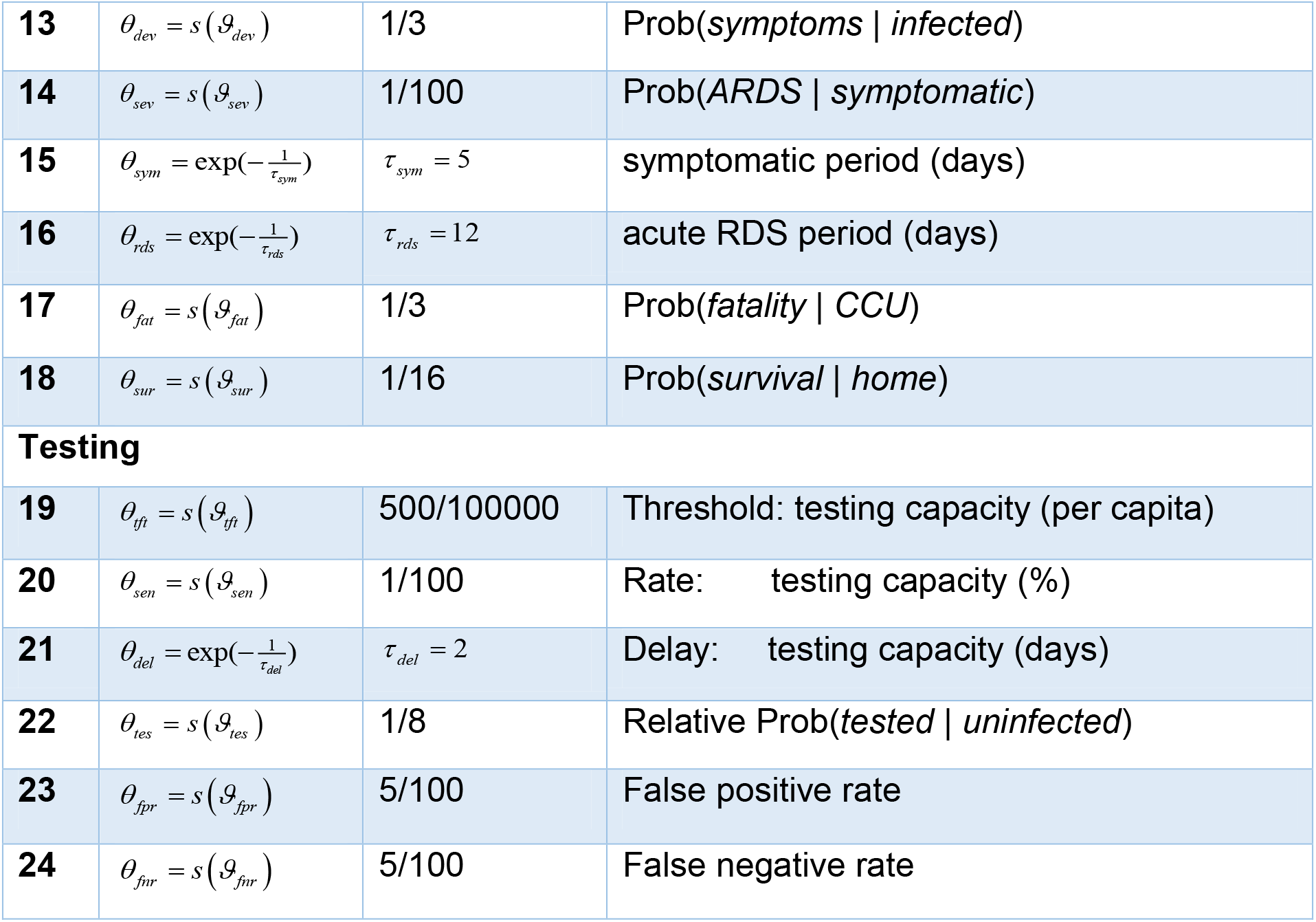
Summary of priors for LIST model parameters. Note that prior variances were fixed to 1 for all unbounded model parameters

At this stage, we note that an important feature of the LIST model is the notion of ‘resistance’. Anecdotal evidence for such resistance comes from, *e*.*g*., the *Charles De Gaulle* warship^6^ and/or the Diamond Princess cruise ship^7^. Similarly, children are just as likely as adults to become infected but are less likely to be symptomatic, develop severe symptoms or become contagious (Bi *et al*., 2020). Such asymptomatic and non contagious people are, under the LIST model, in a ‘resistant’ state of the ‘infection’ factor. The ‘resistant’ state captures possible cross-reactivity mechanisms (Grifoni *et al*., 2020; Ng *et al*., 2020) or other protective host factors (Bunyavanich *et al*., 2020; Zheng *et al*., 2020). This is critical, because it means that the LIST model considers two kinds of immunity: (i) ‘immune’ people, who have acquired immunity by being infected and then contagious (reflected in neutralising antibody titres), and (ii) ‘resistant’ people, who became immune (by other mechanisms *e*.*g*., differential resistance on a genetic basis, cross-immunity, T-cell mediated mechanisms, mucosal-based resistance) without contributing to virus transmission. This form of transmission heterogeneity is partially determined by a specific LIST parameter (*θ*_*res*_, see Equation 7 below), which reflects the proportion of infected people who become resistant.

The distinction between ‘at-home’ and ‘at-work’ people hides another form of transmission heterogeneity. This is because the proportion of ‘at-work’ people determines the number of person-to-person contacts^8^, and hence, the viral transmission speed. Importantly, the LIST model considers social distancing (*e*.*g*., the tendency to stay at home) to be an adaptive feature of epidemic dynamics. In brief, the probability, for a given individual, of going to work (rather than staying at home) is a function of how tight governmental lockdown constraints are and the level of population wide risk-aversion. In the latest derivation of the model (Friston *et al*., 2020c), the former are determined by how close ICU occupancy comes to reach capacity, and the latter is a function of the proportion of symptomatic people in the population. In turn, the probability *P* _→work_ = *P*(*work* | *home, asymptomatic*) of going to work when being asymptomatic is given by:

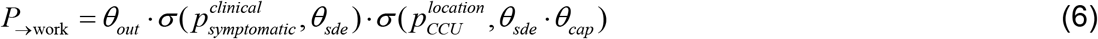

Where *θ*_*out*_, *θ*_*cap*_, and *θ*_*sde*_ are LIST parameters that control the base rate of ‘at-work’ probability, the ICU occupancy capacity and the sensitivity of social distancing to disease symptomatic and ICU occupancy states, respectively. And σ (*x,θ*) is a simple inverse sigmoid function: 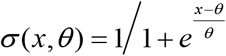.

Figure 6 shows the transition probability *P* _→work_ given in Equation 5, as a function of both the rate 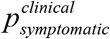 of symptomatic people and the rate 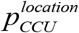 of ICU occupancy, when DCM parameters are set to their prior values.

**Figure 6:**
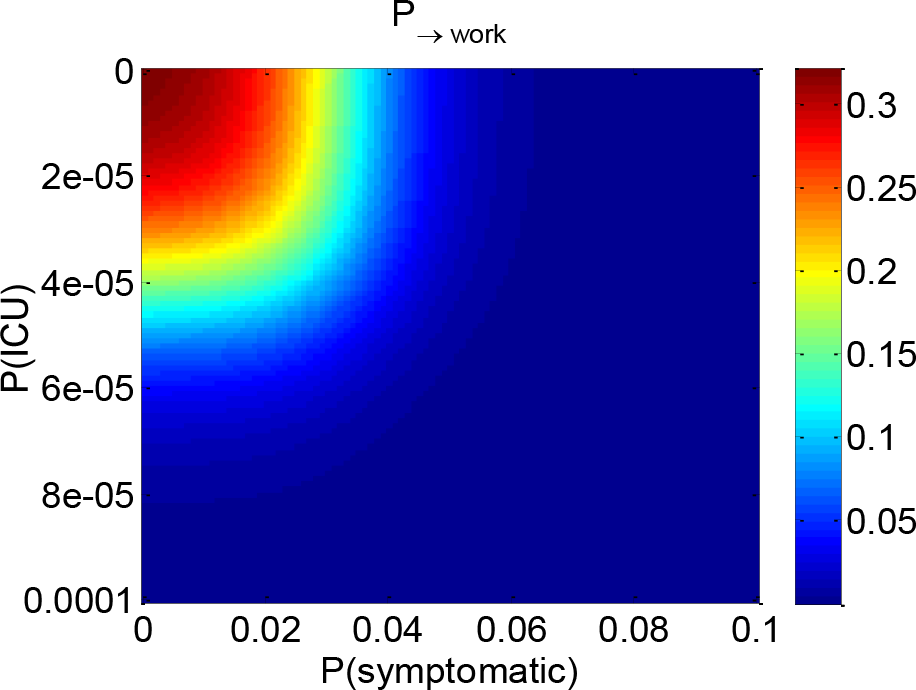
Adaptive social distancing (adapted from Friston *et al*. 2020). The probability *P*_→ work_ (color code) is shown as a function of the rate of symptomatic people (x-axis) and the rate of people located in ICUs (y-axis).

This aspect of the model is important for two reasons. First, it captures the effective adherence to governmental social distancing measures by the population in a flexible way, through its implicit “constraint tightening” parameter *θ*_*sde*_. Second, it can serve to derive an economic cost to the epidemic, in terms of the number of lost working days (see below).

Figure 7 below shows an exemplar simulation of LIST epidemic dynamics, given prior values of model parameters (total duration of simulation is 20 weeks, starting on the day of the first simulated virus infection).

**Figure 7:**
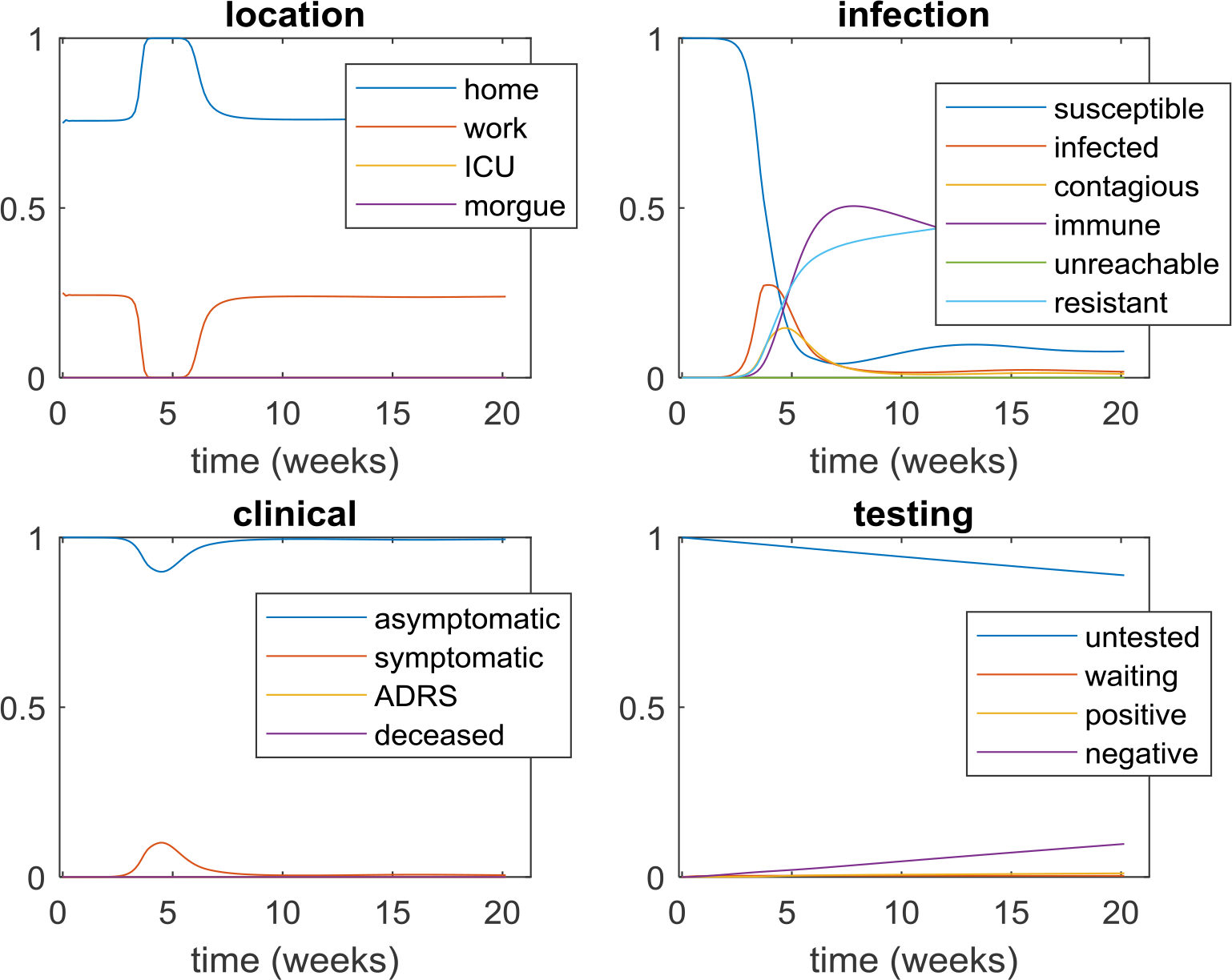
Prior simulation of the LIST model (adapted from Friston *et al*. 2020). Upper-left panel: location factor. Upper-right panel: infection factor. Lower-left panel: clinical factor. Lower-right panel: testing factor. In all panels, the rate of each corresponding state (y-axis, see legends) is plotted as a function of time (x-axis, in weeks). Note that the steady-state probability of being at work (before and after the outbreak peak) is about 1/4. This is because (i) not all people are active and (ii) people typically work 8 hours a day. In addition, one may think that the marginal probability of many LIST barely differs from 0 (*e*.*g*., ICU occupancy or deceased states). This is simply a matter of scaling because these states show a transient increase during the outbreak (which would be visible, would one zoom in). Finally, the ‘unreachable’ state has formally a zero probability here (we will introduce this notion later).

One can see the effect of adaptive social distancing. In brief, as the virus begins to spread, more people become symptomatic and transit to ICUs, which incites people to stay at home during the epidemic outbreak (*cf*. upper-left panel in Figure 7). This prunes the socio-geographical network from some of its ‘at-work’ contacts, which mechanically slows the virus transmission down. Eventually, the pool of susceptible people becomes exhausted, the rate of symptomatic people starts to decrease, and people go back to work.

One can also see a late, slow and small resurgence of susceptible people that peaks about 10 weeks after the main epidemic outbreak (cf. upper-right panel, blue line). This corresponds to an important feedback mechanism that can generate secondary waves: namely, a possible loss of immunity (Friston *et al*., 2020c). As we will see, this mechanism is difficult to evidence from currently available data, because the putative time scale of immunity loss is too slow for observable effects to be present yet. Although this candidate secondary wave-generating mechanism is not induced by lockdown, it may be crucial to consider it in the long run (Friston *et al*., 2020b). Note that, under our prior model parameters, oscillations in the infection factor are too small in magnitude to induce another outbreak that would be visible in terms of, *e*.*g*., an increase in the number of symptomatic carriers. This is because natural resistance to the virus *a priori* attenuates the loss of immunity that is induced by, *e*.*g*., seasonal virus mutations. Importantly, although such a balance between natural resistance and immunity loss remains to be confirmed, we do not expect this kind of COVID reinfection to play a role in the short-term (Bao *et al*., 2020).

In what follows, we consider another candidate second wave generating mechanism, which directly relates to adaptive social distancing.

### b. Lockdown-induced network *unreachability*

In brief, lockdown means that most people stay at home. This does not mean that they cannot be infected (or infect other people). Rather, it means that the number of direct contacts they have with other people is much smaller. It follows that when lockdown becomes effective, some susceptible people have no direct or indirect contact with infected people (*i*.*e*., *via* direct contacts of their own direct contacts). From the perspective of the virus, these people become “*unreachable”*. This is important, because when lockdown ends, people get back to work and become susceptible again, effectively enabling a 2^nd^ wave. Figure 8 below summarizes this lockdown-induced network *unreachability* mechanism.

**Figure 8:**
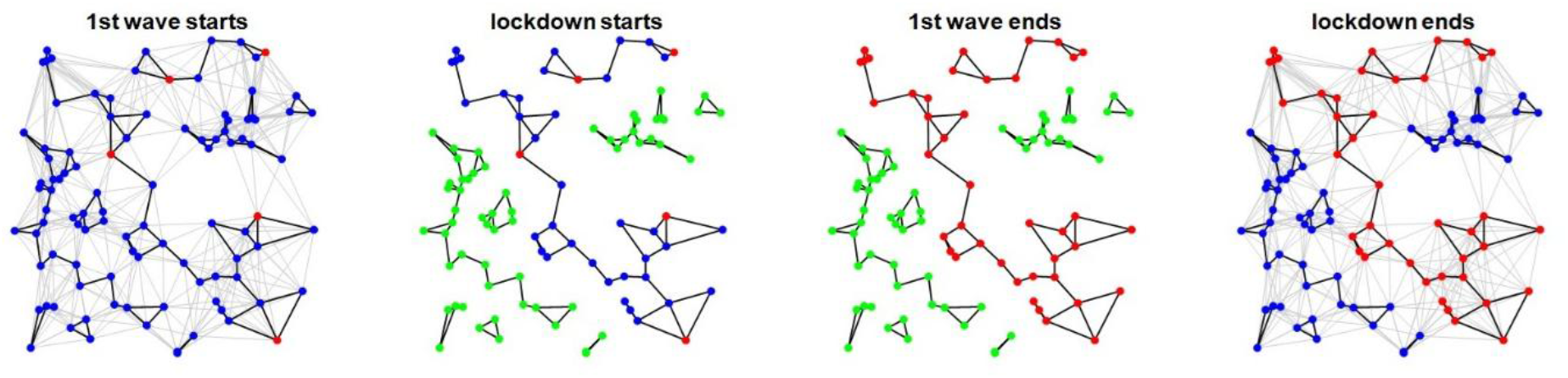
Lockdown-induced network *unreachability*. An example of a socio-geographical network is shown. Black lines depict direct ‘at-home’ contacts, whereas grey lines depict direct ‘at-work’ contacts. Blue, red and green dots show susceptible, infectious and unreachable people, respectively. Before lockdown, all contacts are in place, and few individuals are infected (1^st^ wave starts). When lockdown starts, ‘at-work’ contacts are shut down, and a few people become unreachable because none of their indirect ‘at-home’ contacts are infected. All other susceptible people remain susceptible. While lockdown is in place, the pool of susceptible people becomes exhausted, which signals the end of the 1^st^ wave. At the end of lockdown, ‘at-work’ contacts are switched back on, and unreachable people become susceptible again. This might start a 2^nd^ wave… Note that this example is simplistic, for at least two reasons: (i) not all ‘at-work’ contacts are removed during lockdown (network pruning effectively depends upon adaptive social distancing), and (ii) not all susceptible people may become infected during the first wave (this depends upon the interaction of lockdown duration and biological features of the COVID pandemic such as resistance). We account for these possibilities when modelling network unreachability.

The LIST model already considers the difference between ‘at-home’ and ‘at-work’ contacts. In addition, adaptive social distancing predicts how numbers of people ‘at-home’ and ‘at-work’ will evolve as the virus spreads. What remains to be considered is a feedback mechanism, by which people transit from a ‘susceptible’ to an ‘unreachable’ state, as a function of the marginal rates of both people ‘at-work’ and infected/contagious people, and the average number of ‘at-home’ and ‘at-work’ contacts. The difficulty here lies in the fact that this transition rate depends on the decomposition of the socio-geographic network into *connected components, i*.*e*., clusters of people who are all *indirect* contacts of each other, through direct contacts. The volume of the unreachable compartment will then equal the number of people belonging to connected components that do not include virus carriers.

We therefore conducted Monte-Carlo numerical simulations to assess the functional form of this transition rate. We simulated random socio-geographical networks (network size: 100 nodes) with different average numbers of direct ‘at-work’ (10, 20, 30) and ‘at-home’ (1, 2, 3) contacts, and different proportions of infected people (from 0 to 100%). We also systematically varied the proportions of ‘at-work’ people within the network (from 0 to 40%). We then decomposed the network into connected components^9^ and identified the ensuing rate of unreachable people. The results of these Monte-Carlo simulations are summarized on Figure 9 below.

**Figure 9:**
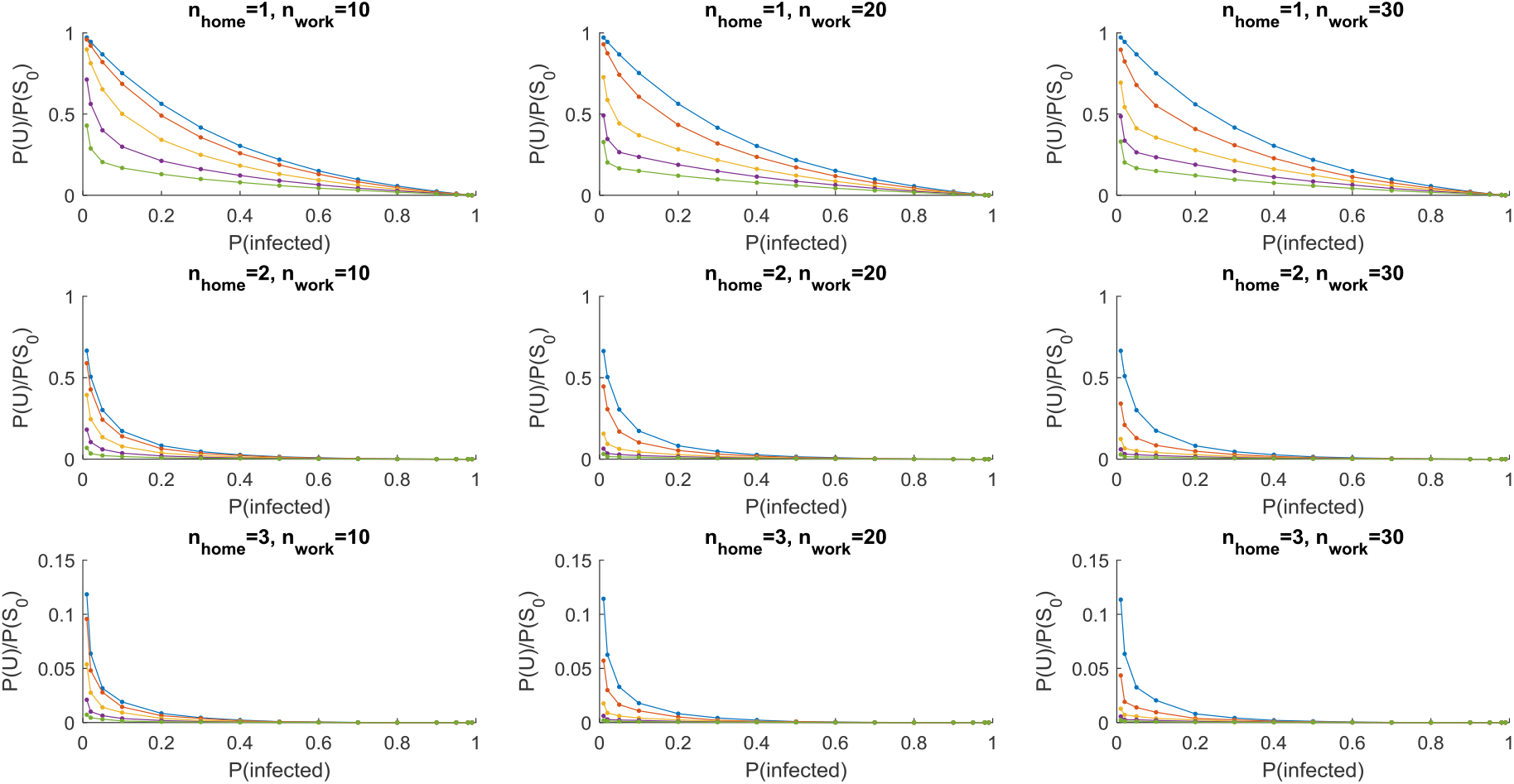
Simulated *unreachability* rate for random networks. In each panel, the ratio P(U)/P(S_0_) of unreachable to initially susceptible people (y-axis) is plotted as a function of the rate of infected people (x-axis) for different numbers of ‘at-work’ people (colour code: blue: 0%, red: 10%, yellow: 20%, violet: 30%, green: 40%^10^). Dots are Monte-Carlo averages over 5,000 simulations. Different panels show the impact of the number of direct ‘at-home’ and ‘at-work’ contacts.

As expected, the unreachability transition rate P(U)/P(S_0_) decreases with the numbers of infected people and those ‘at-work’. Note that when P(infected) tends towards zero, then the rate of unreachability transition tends towards 1 (and reciprocally). But this relationship depends upon the number of people ‘at-work’: when more people are at work, then P(U)/P(S_0_) decreases, everything else being equal. Finally, increasing the number of direct ‘at-home’ contacts further decreases P(U)/P(S_0_).

The precise quantitative aspect of these simulations is unlikely to generalize to non-random socio-geographical networks, with, *e*.*g*. “scale-free” or “small-world” properties (Pastor-Satorras and Vespignani, 2001; Small *et al*., 2006; Xiao Fan Wang and Guanrong Chen, 2003). Nevertheless, we retain the qualitative features of these simulations. The ensuing modifications to the LIST model are described below.

### c. Modified LIST model

Firstly, we include a new ‘unreachable’ state in the ‘infection’ factor. By definition, someone is *unreachable* if he/she does not carry the virus and has neither direct nor indirect contact with virus carriers. In line with the above simulations, the *unreachability* rate is determined by the rates 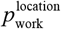 and 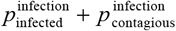of people ‘at-home’ and virus carriers, respectively. This means that the infection factor is now composed of six sub-states: 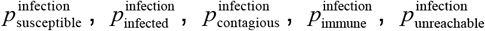 and 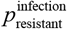.

Secondly, we allow for the possibility that, when susceptible people are at home, they may either stay susceptible, get infected or become unreachable, with a rate that depends upon the proportions of workers and virus carriers. Finally, unreachable people who get back to work (e.g., when social distancing relaxes) may become susceptible again, effectively reversing the *unreachability* flow.

We then modify the conditional transition matrices of infectious states as follows:

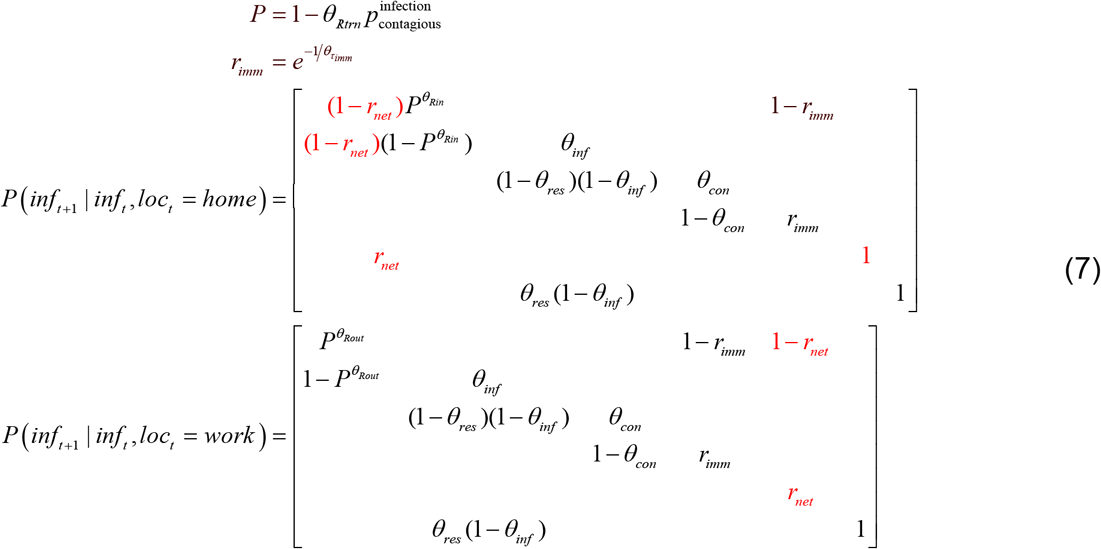

where the *unreachability* rate is given by:

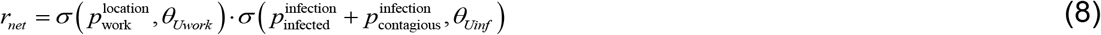

In Equation 8, *θ*_*Uwork*_ and*θ*_*Uinf*_ are unknown LIST model parameters that control the sensitivity of the transition rate to the proportions of ‘at-work’ people and virus carriers, respectively. Prior values for these parameters were obtained by maximizing the match between Equation 8 and simulations on random networks (*cf*. Figure 9). The complete transition matrices of the modified LIST model can be found in the Appendix 1 of this note.

Figure 10 below shows an exemplar simulation of the modified LIST model, given the prior values of the model parameters (as before, total duration of simulation is 20 weeks, starting at the day of the first simulated infected case).

**Figure 10:**
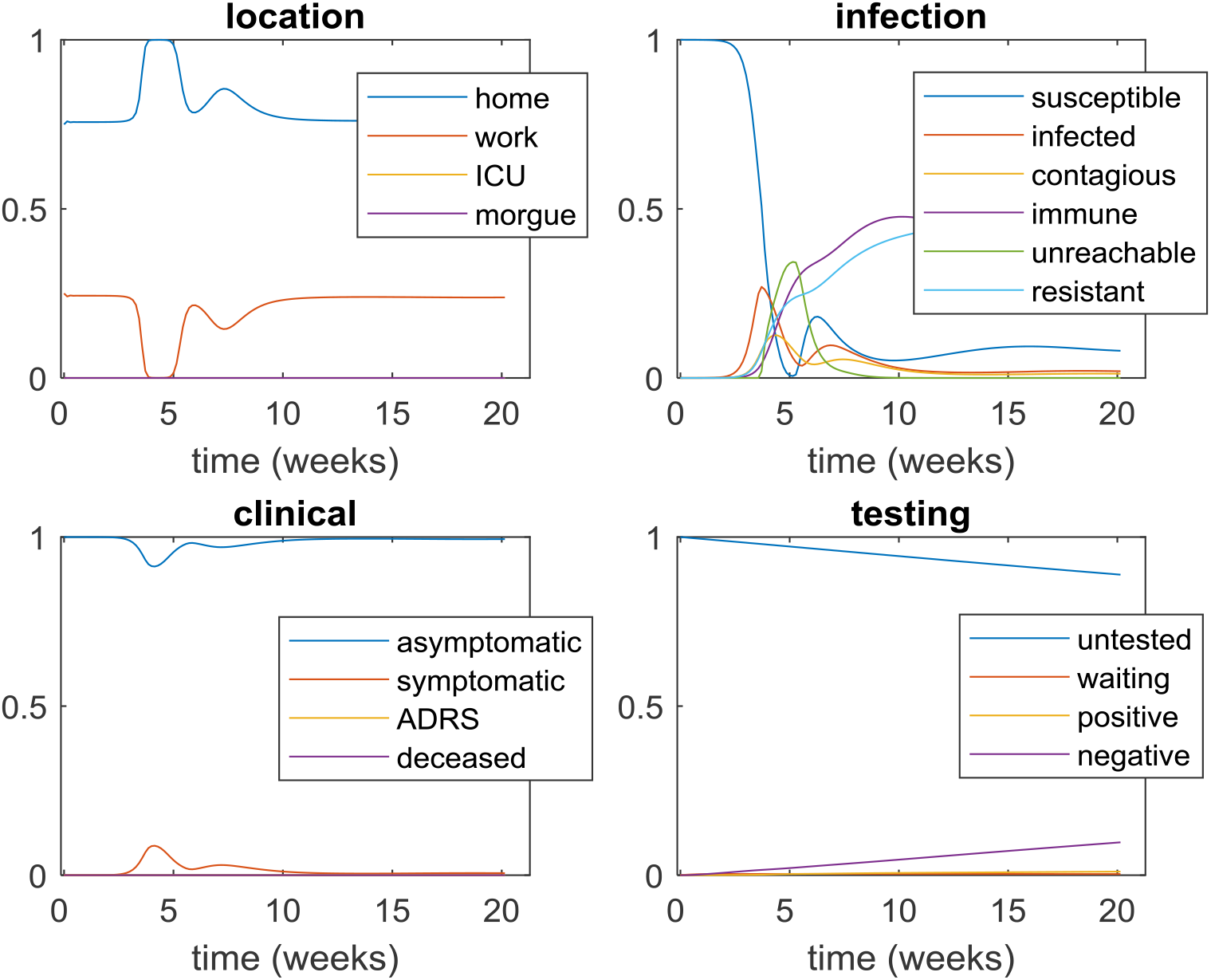
Prior simulation of the modified LIST model. Same format as Figure 7. Note that these simulations are identical to those of Figure 7, except that the modified LIST model now includes transitions to an ‘unreachable’ state in the ‘infection’ factor.

Under the prior model parameters, adaptive social distancing now indirectly induces secondary epidemic outbreaks. In brief, as the virus begins to spread, adaptive social distancing first locks people into their homes, which prunes the network from most of its ‘at-work’ connections. This yields an increase in the rate of unreachable people. Together with virus spreading among the remaining subset of the population, there is eventual exhaustion of the pool of susceptible people, resulting in fewer symptomatic people. In turn, adaptive social distancing relaxes and ‘at-work’ connections are switched back on. This reverses the *unreachability* flow, effectively reinjecting new susceptible people into the network. The latter eventually get infected, which starts another cycle of epidemic outbreak.

In summary, including network *unreachability* within the LIST model may generate secondary waves that peak with a delay of about three weeks after the lockdown offset (defined as the time when people start to go back to work). This timescale is to be compared with the increase of susceptible people 10 weeks after the second outbreak, which is due to loss of immunity. Note that neither Figure 7 nor Figure 10 should be taken as model predictions. They are example simulations of the LIST model, which were chosen to demonstrate LIST mechanisms that can generate secondary waves. In particular, the existence, number, magnitude and delay of secondary waves actually depend upon the interaction of many nonlinear mechanisms (adherence to social distancing, sensitivity of network *unreachability* to proportions of ‘at-work’ people and virus carriers, resistance rate, etc). The effective contribution of these mechanisms is likely to vary from country to country, or even from region to region, and need to be estimated from available epidemiological data. This will be the focus of the next section.

### d. Observable outcomes

Most modelling studies to date rely on daily WHO^11^ or ECDC^12^ data reports, which gather cumulative death and positive test counts across countries. These datasets are made openly available as part of a global collaborative effort to fight the COVID pandemic (see: https://github.com/owid). In addition, a few governmental agencies have recently made an effort to assemble and make openly available richer datasets, including, *e*.*g*., ICU occupancy and remission counts (*e*.*g*., for France see: https://www.data.gouv.fr/fr/datasets/).

As with most modelling studies performed so far during the COVID pandemic, previously communicated applications of the LIST model fitted daily death (hereafter*o*^(1)^) and positive test (*o*^(2)^) numbers only. However, the structure of the model and its associated inversion scheme makes it very easy to add to the generated outcome data with remission (*o*^(3)^) and ICU occupancy (*o*^(4)^) rates:

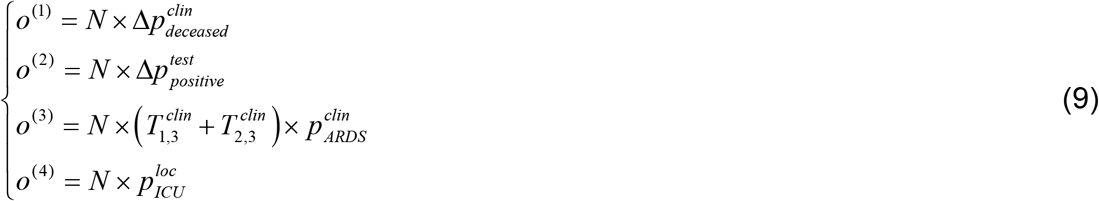

Where 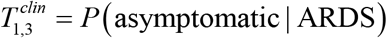 and 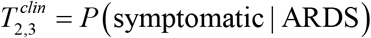 are the daily probability of transiting from ARDS to asymptomatic or (mild) symptomatic states, respectively, and Δ*p ≜ p*_*now*_ − *p*_*yesterday*_ is the daily change in the corresponding marginal probability (with a slight abuse of notation)^13^. The particular form Equation 9 takes for confirmed cases and death numbers derive from the fact that the ‘positive’ and ‘deceased’ states (of ‘testing’ and ‘clinical’ factors, respectively) are so-called “absorbing” states in the LIST model, whereas remission rates and ICU occupancy are not.

Equation 9 shows that observable outcomes provide only partial information regarding internal (latent or hidden) model states. However, in principle, nothing prevents fitting the LIST model to other types of observable data, such as measures of population adherence to social distancing and/or serological surveys. At the time of writing, no other data types were reliably and regularly accessible.

## 4. Results: analysis of regional French data

### a. Dataset description and analysis plan

We extracted four reported time series of daily deaths, positive cases, people in remission and ICU occupancies in all French metropolitan regions (*Auvergne-Rhône-Alpes, Bourgogne-Franche-Comté, Bretagne, Centre-Val-de-Loire, Grand-Est, Hauts-de-France, Normandie, Nouvelle-Aquitaine, Occitanie, Pays-de-la-Loire, Provence-Alpes-Cote d’Azur* and *Ile-de-France*) from *Santé Publique France*^14^. These time series start on the 11th of March and end on the 6th of June 2020. We first interpolated missing data, and then smoothed the resulting time series with a gaussian kernel of 4 days FWHM^15^. To estimate region-specific model parameters, we relied on the variational Laplace approach to approximate Bayesian inference (Daunizeau, 2018; Friston *et al*., 2007) that is implemented in VBA academic freeware (Daunizeau *et al*., 2014). Note that the modified LIST model was fitted to the 4 timeseries concurrently. We then extrapolated the ensuing LIST hidden states to derive predictions until the 31th of December 2020. We also derived counterfactual predictions of what would have happened, had adaptive social distancing been inactive. These analyses serve to highlight what the alternative scenario of the LIST model implies, in terms of the public health benefit and the economic cost of lockdown.

### b. National level summary of LIST dynamics

To begin with, we summarize the predicted dynamics of observable outcomes, *i*.*e*., daily death rates, positive case rates, remission rates and ICU occupancy. We derived estimates of these quantities at the national level simply by summing the model-based predictions over regions. The resulting dynamics cover the full 2020 year and are shown on Figure 11 below.

**Figure 11:**
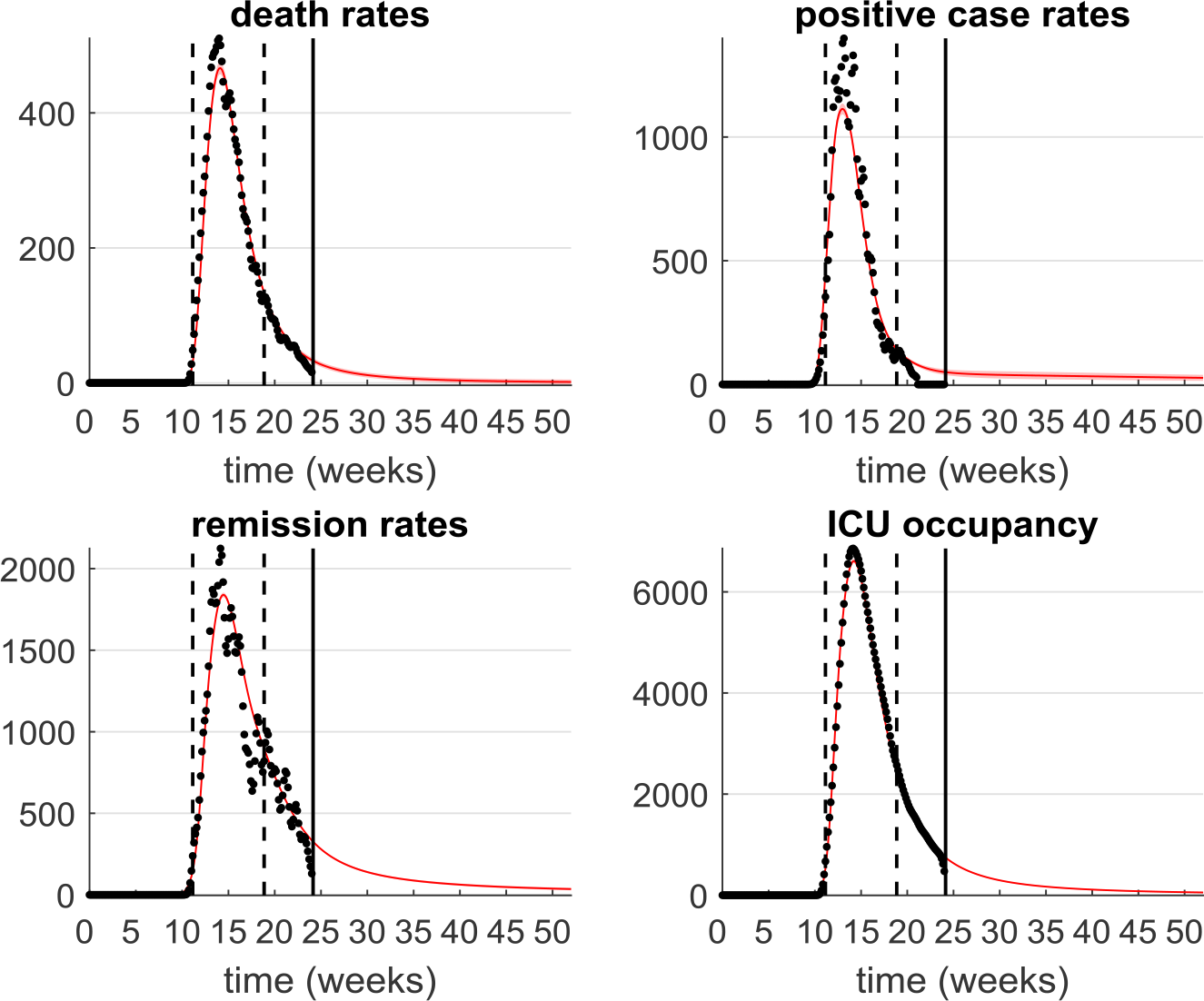
Fitted and predicted outcomes at the French national level. In all panels, plain red lines show the predicted outcome (along with its standard deviation across regions), and black dots show the observed (interpolated and smoothed) outcomes as a function of time (x-axis, in weeks, from the 1st of January to the 31st of December 2020). The two dashed black lines show the start and end of governmental lockdown. The solid black line is the 17th of June 2020 (date of writing of this manuscript). Upper-left panel: daily death rates (only hospital data). Upper-right panel: daily positive case rates (only laboratory data). Lower-left panel: daily remission rates. Lower-right panel: daily ICU occupancy.

Reassuringly, all observable outcomes are accurately fitted, *i*.*e*. the LIST model accurately captures the current outbreak of the epidemic. Interestingly, all predicted outcome dynamics show a rapid growth rate around the onset of governmental lockdown and a remarkably slow decay rate to steady state. Importantly, no post-lockdown secondary outbreak is visible (yet). But why did no secondary waves appear? And how does this pre-to post-lockdown asymmetry come about? As we will see below, the explanation (under the LIST model) involves both network *unreachability* and resistance mechanisms.

We now report the latent causes of the above outcomes; namely, the estimated dynamics of the location, infection, clinical and testing factors at the national level. Recall that, having fitted and extrapolated the LIST model, one obtains daily estimates of the region-specific marginal probabilities of being in each state of each LIST factor from the 1st of January 2020 to the 31th of December 2020. We then derive national estimates from a weighted average of the region-specific dynamics where the weights are the relative population sizes of each region. The ensuing national estimates of LIST hidden state dynamics are summarized on Figure 12 below.

**Figure 12:**
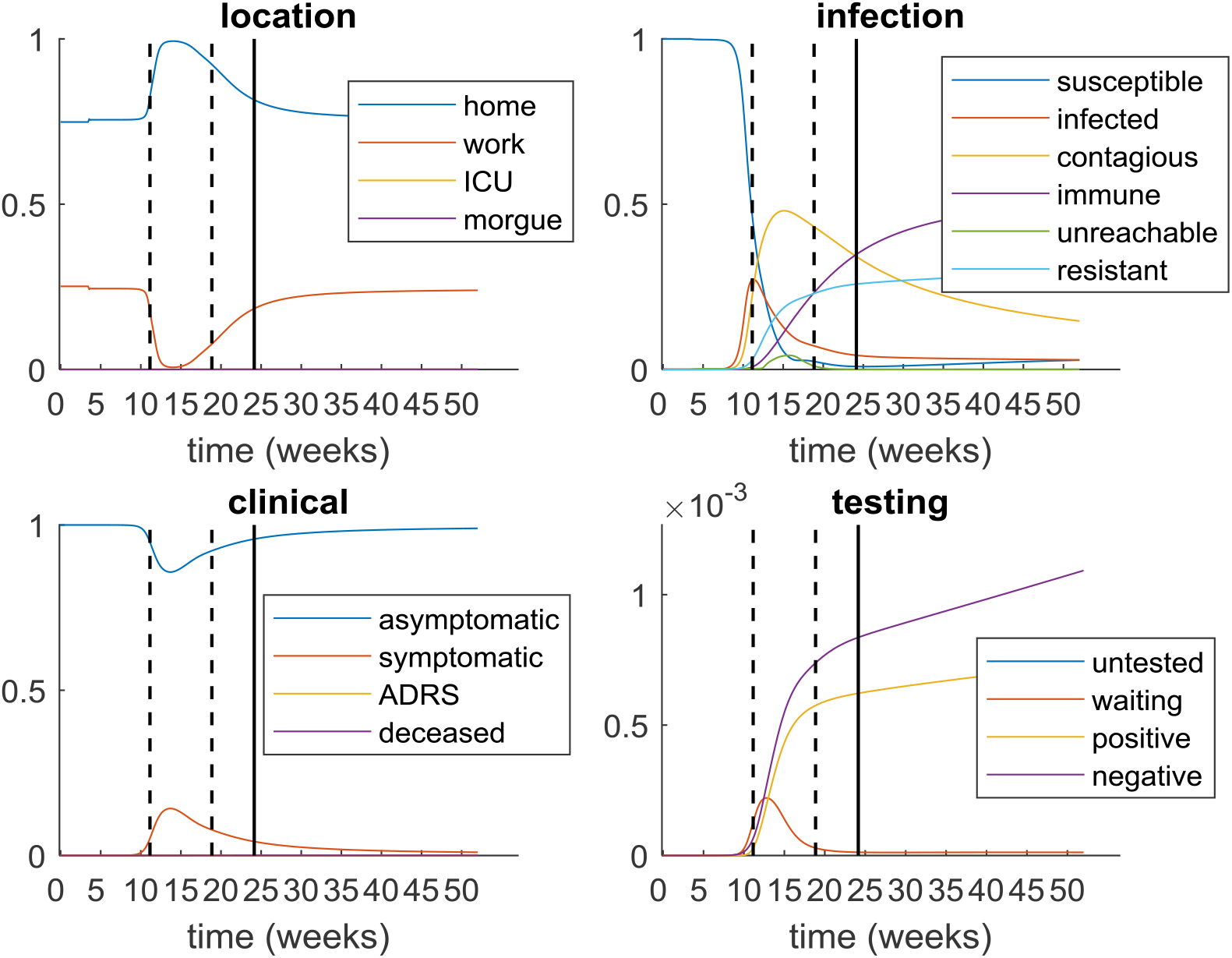
Estimated LIST dynamics at the French national level. In all panels, the marginal probability of being in each corresponding state (y-axis) is shown as a function of time (x-axis, in weeks, from the 1st of January to the 31st of December 2020). Upper-left panel: location factor. Upper-right panel: infection factor. Lower-left panel: clinical factor. Lower-right panel: testing factor (here, we zoomed around the origin of the y-axis to disclose the - otherwise insignificant - testing dynamics).

First, note that the estimated location factor hidden states show a stiff change at the start of the governmental lockdown. Thus, according to the LIST model, most people effectively stayed at home during lockdown, and then slowly started to go back to work. This is remarkable, given that the LIST model was not informed at all about governmental lockdown policy. At the time of writing, we estimate that about two thirds of the active French population have returned to the workplace. Therefore, this provides face validity to the LIST model inference on adaptive social distancing. Second, symptomatic cases (‘clinical’ factor) show the same protracted dynamics as above: namely a temporary stiff increase and then a slow decay back to steady state. The number of virus carriers (*i*.*e*., infected and contagious people from the ‘infection’ factor) also evolves with similar dynamics. There is no strong evidence for lockdown-induced secondary waves, despite a visible transient increase in the rate of unreachable people (cf. green line in the upper-right panel of Figure 12). In brief, although people adhered to social distancing, this effectively slowed the post-lockdown epidemic dynamics, without generating secondary outbreaks.

Also, at the onset of governmental lockdown, about 50% of the population was carrying the virus. And, by the time lockdown ended, 45% of the population was still contagious but about 40% had acquired immunity (half from natural resistance to the virus). Looking at the infection rate more closely, it appears that the peak in the number of infected people occurred just before lockdown onset. Having said this, there may have been little reason to start the lockdown sooner because, according to the LIST model, most virus carriers were asymptomatic at that time. Note that such high rates of asymptomatic virus carriers is in line with recent biomedical surveys (Lavezzo *et al*., 2020).

In brief, rather than generating catastrophic secondary outbreaks (as is typically assumed), the impact of lockdown-induced variations in population susceptibility and transmission seems to reduce to a steady-state endemic equilibrium with a low but stable infection rate. Now, this short summary at the national level needs to be nuanced at the regional level. We next inspected between-region variability, which expresses itself in terms of the magnitude and timing of observable epidemiologic outcomes. As we will see, we find evidence that secondary outbreak waves have not happened *despite* strong lockdown-induced variations in social distancing and their ensuing network *unreachability* rate changes.

### c. Between-region variability

Figure 13 below summarizes the observable between-region variability, in terms of variations in post-lockdown mortality rate and prevalence^16^ at the time of writing.

**Figure 13:**
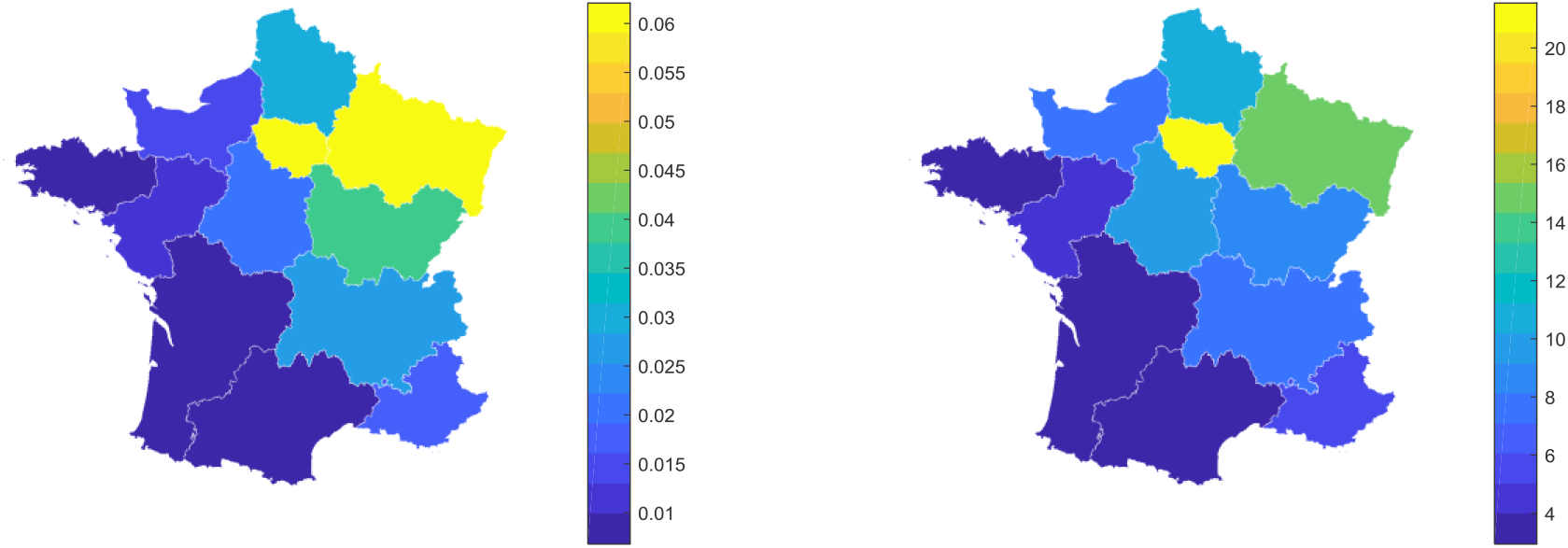
Mortality rates and prevalence across French regions. The mortality rate (left panel, in % of the population size) and prevalence (right panel, in % of the population size) at the offset of governmental lockdown for all French regions. We note that regional prevalence based upon COVID test reports are likely to be strongly under-estimated, because (i) COVID tests have poor sensitivity^17^(Arevalo-Rodriguez et al., 2020; Watson et al., 2020) and (ii) mostly symptomatic people are tested (Gandhi et al., 2020). In fact, LIST estimates of the proportion of virus carriers in the French population are higher than these simple data-driven estimates (despite the slight underestimation of positive test rates that can be seen on Figure 11, upper-right panel).

There is considerable between-region variability in the observed mortality and prevalence figures. Under the LIST model, observed prevalence and mortality are determined by nonlinear interactions between opponent mechanisms (including heterogeneous susceptibility and transmission). In turn, most of the observed between-region variability may result from differences in the phasing of regional LIST ‘infection’ states dynamics with respect to lockdown. This motivated a region-specific analysis of LIST dynamics, which we summarize below.

Beforehand, note that a simple summary statistic of the region-to-region LIST fit accuracy is presented in Appendix 2. In brief, the LIST model explains about 95% of the variance in the 4 observable outcomes (averaged over all regional timeseries).

First, we asked what drives between-region variability in current mortality rates. Recall that network *unreachability* captures a temporary protection, which lasts until adaptive social distancing is relaxed. Therefore, we reasoned that the rate of *unreachability* during lockdown should determine current mortality rates. Figure 14 below tests this prediction.

**Figure 14:**
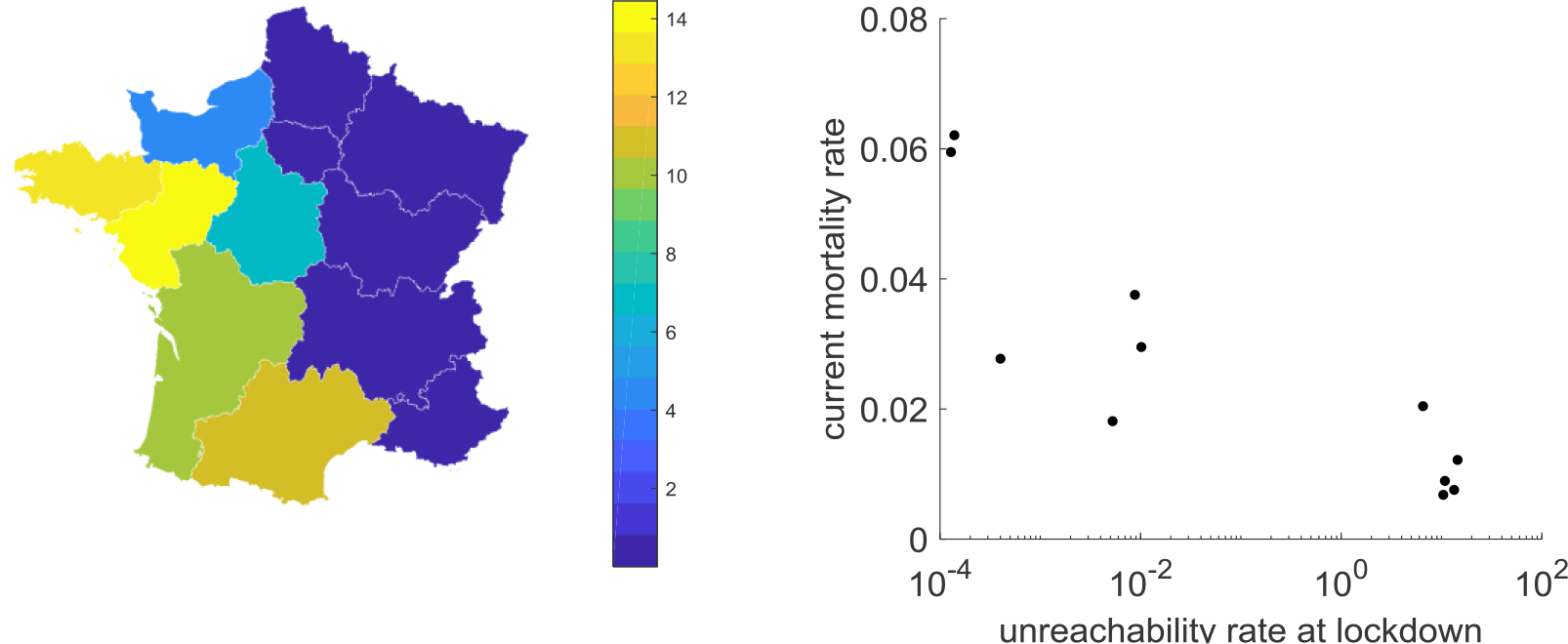
Network *unreachability* rates and mortality rates. Left panel: estimated unreachability rates (in % of the population size) at the middle of the governmental lockdown are shown for all French regions. Right panel: The current mortality rate (y-axis, in % of the population size) is plotted as a function of the unreachability rate at the middle of the governmental lockdown (x-axis, in % of the population size, log scale).

There is considerable variability in the estimated *unreachability* rate in the middle of governmental lockdown (from about 1% in eastern regions to 14% in *Loire-Atlantique*). A simple explanation here is that, by the time lockdown started at the national level, many people were already infected (though most likely asymptomatic) in eastern regions. This means that adaptive social distancing may have had very little impact on virus propagation in eastern regions. In turn, there was little potential for lockdown-induced secondary outbreaks in these regions. This may not have been the case for western regions, whose infection rates began to rise later. Importantly, the current mortality rate clearly decreases as *unreachability* numbers increase. This means that some of the current between-region variability in mortality rates will disappear once the pools of unreachable people (in western regions) become susceptible again and then become infected.

What drives between-region variability in current prevalence? Of particular importance is the resistance rate, which reduces the pool of virus carriers in the early stages of infection. Therefore, we reasoned that current prevalence should decrease when resistance rates (during lockdown) increase. Figure 15 below tests this prediction.

**Figure 15:**
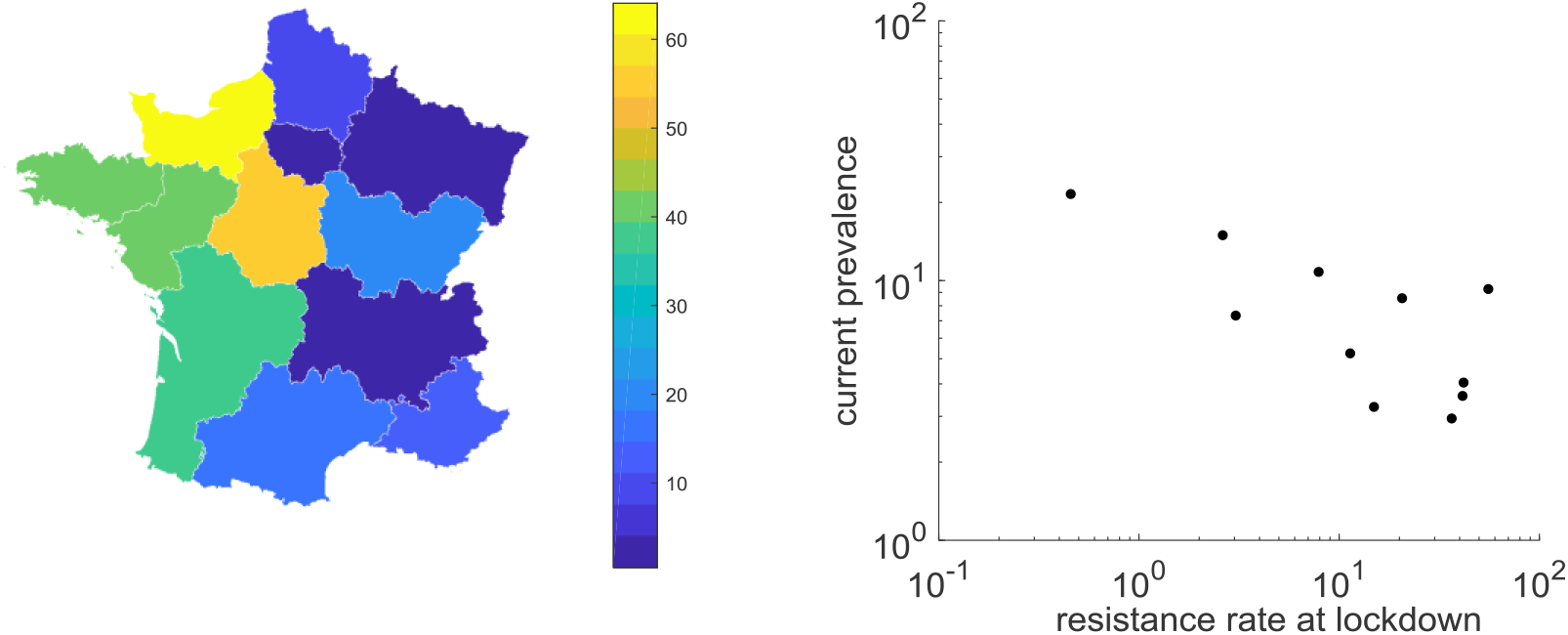
Resistance rates and prevalence. Left panel: resistance rates (in % of the population size) at the middle of the governmental lockdown are shown for all French regions. Right panel: The current prevalence (y-axis, in % of the population size, log-scale) is plotted as a function of the resistance rate at the middle of the governmental lockdown (x-axis, in % of the population size, log scale).

Here again our reasoning holds. Additionally, it is clear that the resistance rate is much higher in western than in eastern regions. Note that, depending on the phasing of local infection dynamics w.r.t. lockdown, network *unreachability* and resistance may act as opponent mechanisms. This implies that the observed relationship between current prevalence and resistance may non-trivially depend upon adpative social distancing. In fact, both current estimates of mortality and prevalence are impacted by resistance and *unreachability* rates over time.

At this point, it is important to note that under the LIST model, regions may differ greatly in terms of the dynamics of their respective epidemic outbreak. For example, population adherence to social distancing may be heterogeneous. In turn, regions where the *unreachability* rate (during lockdown) was high will have slower rates of virus propagation, and hence higher numbers of virus carriers (by the end of this year). In addition, regions may differ in terms of the rate at which immunity is lost (Friston *et al*., 2020b). Finally, regions where resistance rates are high will have lower numbers of contagious people over time, which may have opposing effects on future immunity levels. Thus, one may ask what level of immunity (composed of both ‘immune’ and ‘resistant’ states in the LIST model) will be acquired in distinct regions. This is summarized in Figure 16 below. Note that this level of immunity includes resistant subpopulations that have not seroconverted. In other words, one would expect the seroprevalence to be much less than the estimates in Figure 16.

**Figure 16:**
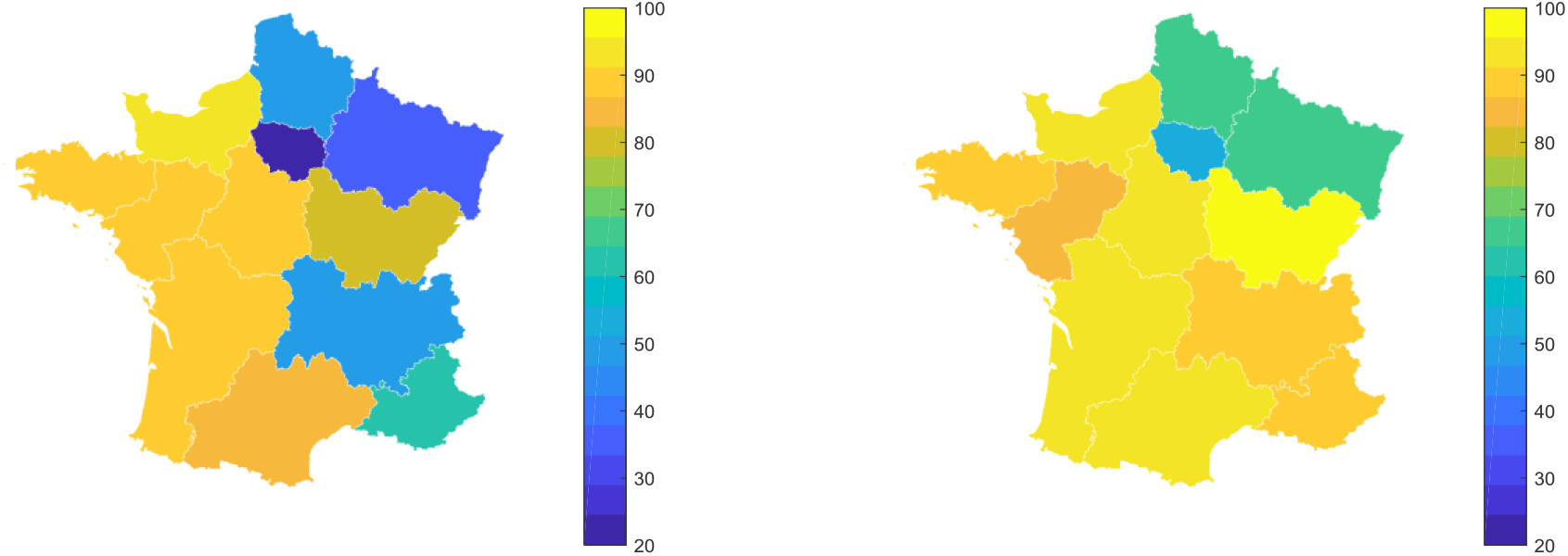
Current and predicted immunity levels. Left panel: current immunity levels (‘immune’ plus ‘resistant’ rates, in % of the population size) at the time of writing are shown for all French regions. Right panel: predicted immunity levels (in % of the population size) at the end of December 2020.

Both current and predicted immunity levels vary greatly across regions: from 20% in *Ile-de-France* at the time of writing to about 90% in all western and southern French regions by the end of December 2020. This variability originates from differences in the proportions of resistant and infected people (which peak at different times for different regions). In any case, this has clear implications for current and/or future serological surveys of the COVID pandemic. We note that the rate at which immunity is acquired (difference between predicted and current immunity levels) is mostly controlled by *unreachability* rates during lockdown (results not shown).

### d. Assessing the impact of lockdown

Finally, we exploit the ability to explore model-based counterfactual scenarios. Having fitted the LIST model to regional data, we ask what would have happened if there had been no adaptive social distancing, and hence no governmental lockdown. Here, at least two outcomes are of interest. In terms of public health, how many people would have died (up to now)? Conversely, we quantify how many lives were saved with social distancing. Under the SIR model, the health impact of lockdown has to be high, because social distancing is, by assumption, the reason why the rate of infected people eventually declined, despite the “fact” that herd immunity has not been reached. However, the LIST model suggests that the infection and resistance rates at the onset of the lockdown were much higher than SIR estimates. As we will see, this implies that the number of saved lives may be much lower than originally thought^18^. In terms of economics, we calculate how many working days were lost (up to now). Under the LIST model, this number is directly derived from comparing the cumulated numbers of people ‘at work’ with and without adaptive social distancing^19^. These outcomes are summarized in Figure 17 below. Note that, to enable a viable region-to-region comparison, we normalize these indices by population size in each region.

**Figure 17:**
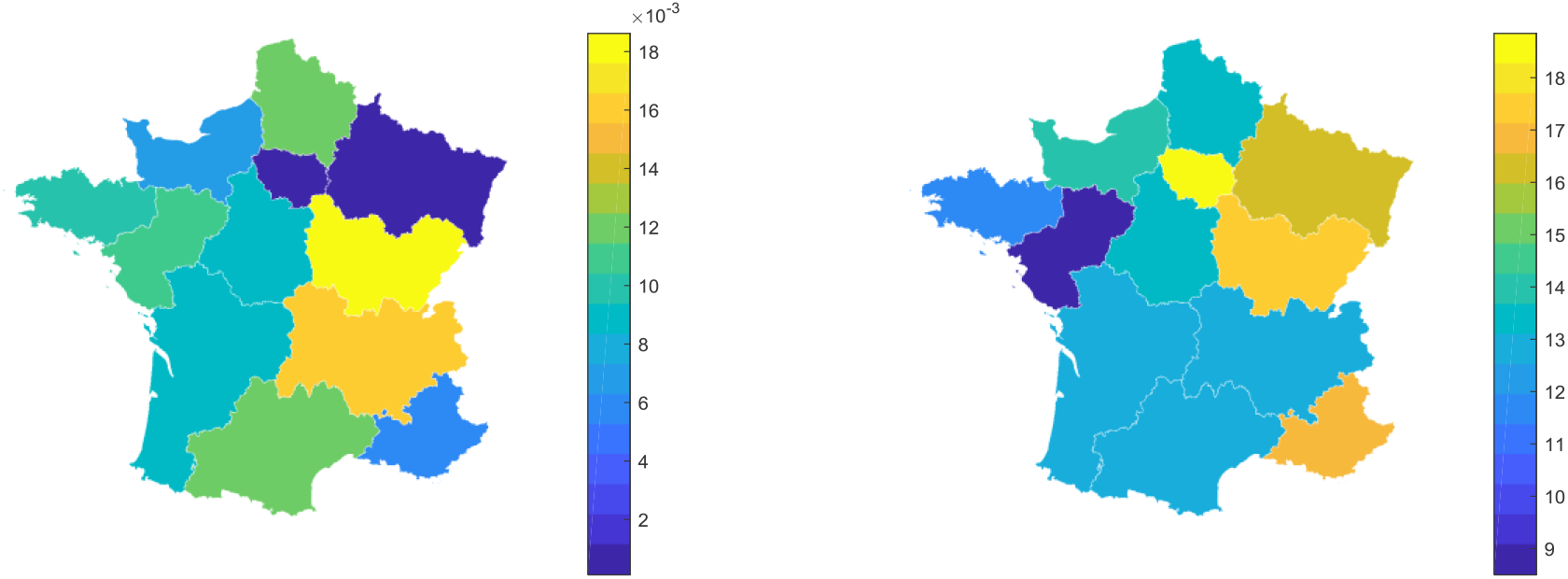
Saved lives and lost working days. Left panel: the number of saved lives (up to now, per people) is shown for all French regions. Right panel: the number of lost working days (up to now, per people) is shown.

Both health and economic outcomes vary greatly across regions. Importantly, regions where most lives were saved are not necessarily those where most working days were lost. Consider *Bourgogne-Franche-Comté*, for example. This is the region where, according to the LIST model, most lives were saved (relative to its population size). In absolute terms, this corresponds to 518 lives saved, for a population of about 2,783,000 people (to be compared with 1030 hospital deaths). In this region, about 17 working days were lost per inhabitant, which is almost identical to *Provence-Alpes-Cote d’Azur*. However, “only” 303 lives were saved for 5,055,000 inhabitants in that region (to be compared with 923 hospital deaths). These differences arise from a combination of biological (*e*.*g*. natural resistance rates) and demographical (*e*.*g*. social distancing adherence) factors. This result implies that, in principle, the unavoidable cost-benefit trade-off of governmental lockdown measures may be improved by adopting adaptive local policies that account for inter-regional differences.

## 5. Discussion

In this work, we have extended the LIST model to account for the potential impact of adaptive social distancing on the generation of secondary epidemic waves. In particular, we focused on the notion of network *unreachability, i*.*e*., the fact that when ‘at-work’ connections get pruned from the network, some people may no longer be susceptible to the virus. We have demonstrated the impact of this mechanism on the COVID epidemic using both numerical simulations and data analyses. In particular, we have shown that, rather than generating catastrophic post-lockdown secondary outbreaks, this type of process interacts with other nonlinear mechanisms, eventually yielding protracted post-lockdown dynamics of epidemiological outcomes. We have also shown that observable inter-regional differences, including the impact of governmental lockdown on health and economic outcomes, may be partially explained by regional idiosyncrasies in resistance and *unreachability* rates (which themselves may partially originate from differences in the relative timing of infection dynamics *w*.*r*.*t*. lockdown).

For the purpose of our demonstration, we made a number of predictions regarding epidemiological outcomes, including death rates and acquired immunity levels. At this point, we acknowledge a few weaknesses of our approach.

First, we derived our predictions from model-based analyses of available epidemiological French data: namely, daily death rates, positive case rates, ICU occupancy and remission rates. These time series are already more informative than most data available from international data repositories (which only report death and positive case rates). Nevertheless, currently available data are noisy (cf. weekly artefactual drops, heterogeneous sampling by testing laboratories, *etc*.), preliminary (from the perspective of the global time course of the pandemic), and scarce (they do not relate to all hidden factors of the LIST model). In particular, should time-resolved data regarding population adherence to social distancing and/or immunity levels be available, the reliability of model-based predictions will be considerably strengthened (Daunizeau *et al*., 2020). Organizing systematic reporting of these types of data during this or future epidemics should be a matter of high priority for quantitative epidemiology.

Second, we did not evaluate the sensitivity of model-based predictions to prior constraints on unknown model parameters. This, in fact, is directly related to the previous comment, because if the data were sufficiently informative, then the inference would not depend upon priors. Having said this, we do not claim that our predictions are more (or less) reliable than those derived from established epidemiologic models (Kissler *et al*., 2020; Lin *et al*., 2020; Salje *et al*., 2020). In fact, most recent modelling studies do not even consider the possibility of challenging their prior estimates. Rather, they either simply fix parameters or work out best-case/worst-case scenarios that derive from arbitrary variations in key transition parameters. Only retrospectively, once the global pandemic ends, will evaluation of the accuracy of model-based predictions and/or performance by solid statistical model comparisons become possible.

We note that the analyses we report here are far from exhaustive. For example, we did not assess the precise phasing of regional LIST infection sub-states dynamics w.r.t. to lockdown and related social distancing measures (whose monitoring at the national level is available from, e.g., ECDC database repositories). This would have enabled a region-by-region quantification of regional secondary outbreak likelihoods. We will pursue this in subsequent publications. Also, many important biodynamical properties of COVID infection, which one would ideally want to include in LIST-like models, are still unknown. This is important because, although a lockdown-induced secondary wave is unlikely (at the national level), other mechanisms may generate secondary COVID outbreaks. For example, long-term immunity losses, as induced by, e.g., virus mutations, may eventually cause a resurgence of infection rates (Friston et al., 2020b). Because of their slow timescale, such mechanisms are difficult to evidence from the currently available data. Therefore, our conclusions regarding the possible endemic state of the French epidemic would need to be nuanced, if/when reliable data regarding candidate processes of immunity loss become available (Edridge et al., 2020; Prompetchara et al., 2020).

All things considered; our main contribution is to provide a computational framework that is efficient enough to support a complexity of epidemiological models that is fit for purpose. On the one hand, using simple SIR-like models that have few parameters may yield intuitive predictions, but may not be complex enough to predict critical epidemiological features (such as secondary waves) that arise from nonlinear dynamical mechanisms. On the other hand, including as many relevant processes as possible in epidemiological models may turn out to be computationally unfeasible, or statistically inefficient. This point is reminiscent of computational modelling in neuroscience, where very similar trade-offs are intensely debated (Daunizeau *et al*., 2011; Eliasmith and Trujillo, 2014; Friston, 2012; Friston *et al*., 2017; Lebreton *et al*., 2019; Palminteri *et al*., 2017). In line with an all-too-famous quote, we tend to think that everything should be made as simple as possible, but no simpler. In the context of COVID pandemics, we would advocate for (i) considering as many sources of heterogeneous susceptibility and transmission as possible, and (ii) relying on solid and efficient approximate statistical inference approaches. It seems to us that the former is required to derive (non-intuitive) predictions regarding future immunity levels and/or secondary waves. The corollaries are that epidemiological models will need to encompass feedback nonlinearities, and risk becoming computationally demanding, which motivates the latter. As in computational neuroscience, our code (including the LIST model and the associated variational Bayesian inference approach) is made openly available from academic freeware repositories^20^.

## Data Availability

Data are available upon request to the corresponding author of this manuscript.

## Appendix 1 the LIST model

Below is a brief summary of the LIST model, as defined in terms of conditional transition matrices of LIST states.

First, recall that the LIST model is defined in terms of the following factorial state-space:

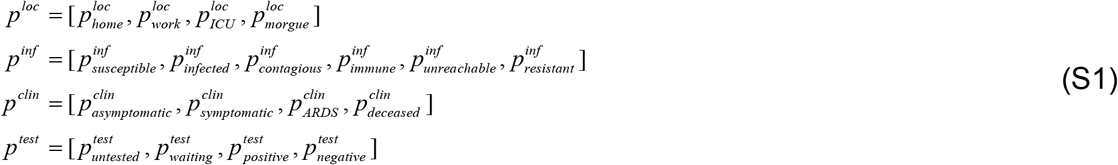

Note that the order of states in each factor matters because it determines the column/row indices of transition matrices.

First, the location factor evolves according to the following transition probability matrices (conditional upon the infection status factor):

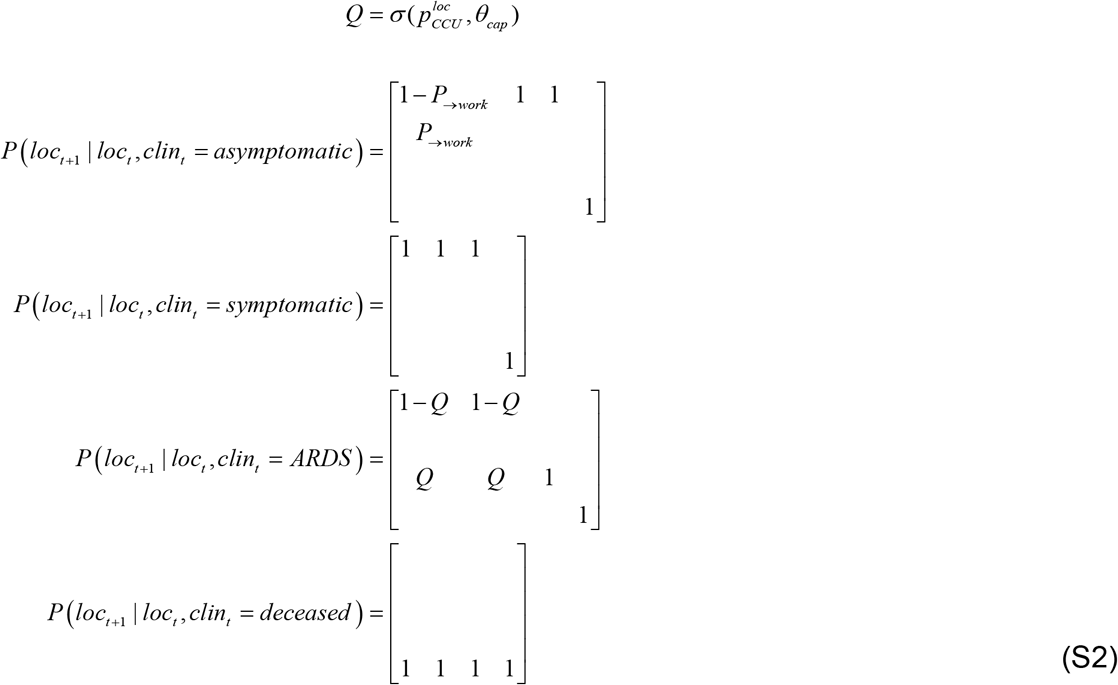

where *P* _→work_ ≜ *P*(*loc*_*t*+2 = *work*_ | *loc*_*t*_ = *home, clin*_*t*_ = *asymptomatic*) is the probability of going to work when asymptomatic (see Equation 6 of the main text). Equation S2 essentially states that (i) if am asymptomatic, I will go to work with a probability *P*_→ work_ (see main text), (ii) if I am symptomatic, I will go back home, and (iii) if I suffer from ARDS, I will move to an ICU with a probability *Q* that is modelled as a sigmoid function of ICU occupancy.

Second, the infection status factor evolves according to the following transition probability matrices (conditional upon the location factor):

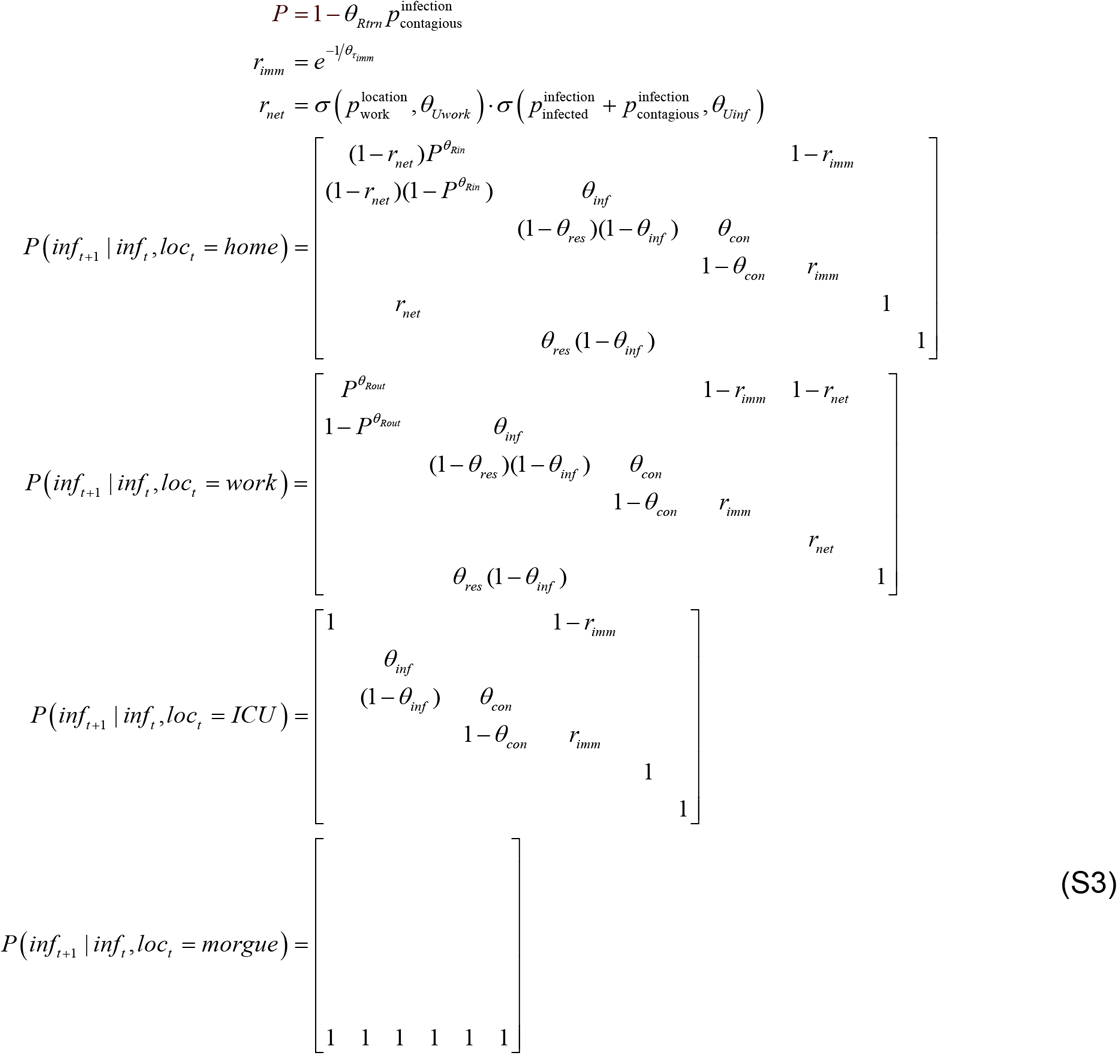

Equation S3 formally completes Equations 7-8 of the main text. When susceptible, one can either remain susceptible, become infected or become unreachable. Note that the latter is only possible when at home, and with a transition rate *r*_*net*_ that depends upon the proportions of ‘at work’ and infected people (see Equation 8 of the main text). If a susceptible individual does not become unreachable, then the probability of becoming infected depends upon the number of social contacts—which depends upon the proportion of time spent at home. This dependency is parameterised in terms of a transition probability per contact (*θ*_*trn*_) and the expected number of contacts at home (*θ*_*Rin*_) and work (*θ*_*Rout*_). Once infected, one remains in this state for a period of time that is parameterised by a transition rate (*θ*_*inf*_). Then, a fraction *θ*_*res*_ of people become resistant and the rest become contagious, before acquiring immunity (with a rate 1-*θ*_*con*_). People can then lose their immunity and become susceptible again at a rate 1-*r*_*imm*_. Finally, the *unreachability* flow can be reversed (from unreachable to susceptible) when at work, with a transition rate 1-*r*_*net*_.

Third, the clinical/symptom status factor evolves according to the following transition probability matrices (conditional upon the infection status factor):

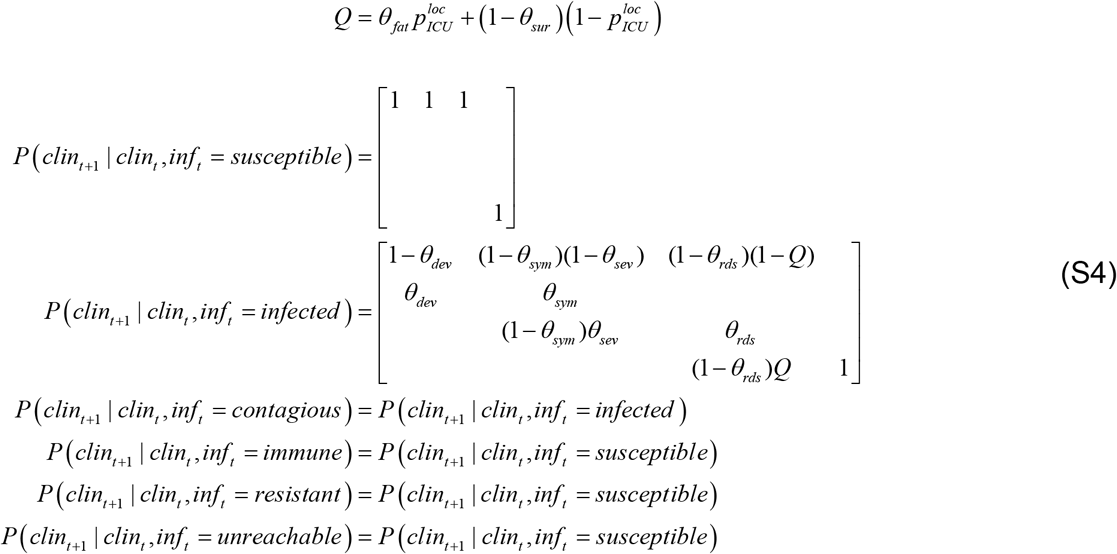

If I am not infected, then I will move to the asymptomatic state, unless I am dead. However, if I am infected, I will develop symptoms with a probability *θ*_*dev*_. Once I have developed symptoms, I will remain symptomatic with a probability *θ*_*sym*_, and either develop ARDS with a probability *θ*_*sev*_ or recover to an asymptomatic state. The parameterisation of these transitions depends upon the typical length of time that I remain symptomatic (*θ*_*sym*_); similarly, when in acute respiratory distress (*θ*_*rds*_). However, I may die following ARDS, with a probability *Q* that depends upon whether I am in an ICU, or elsewhere (*e*.*g*., at home).

Fourth, the testing status factor evolves according to the following transition probability matrices (conditional upon the infection status factor):

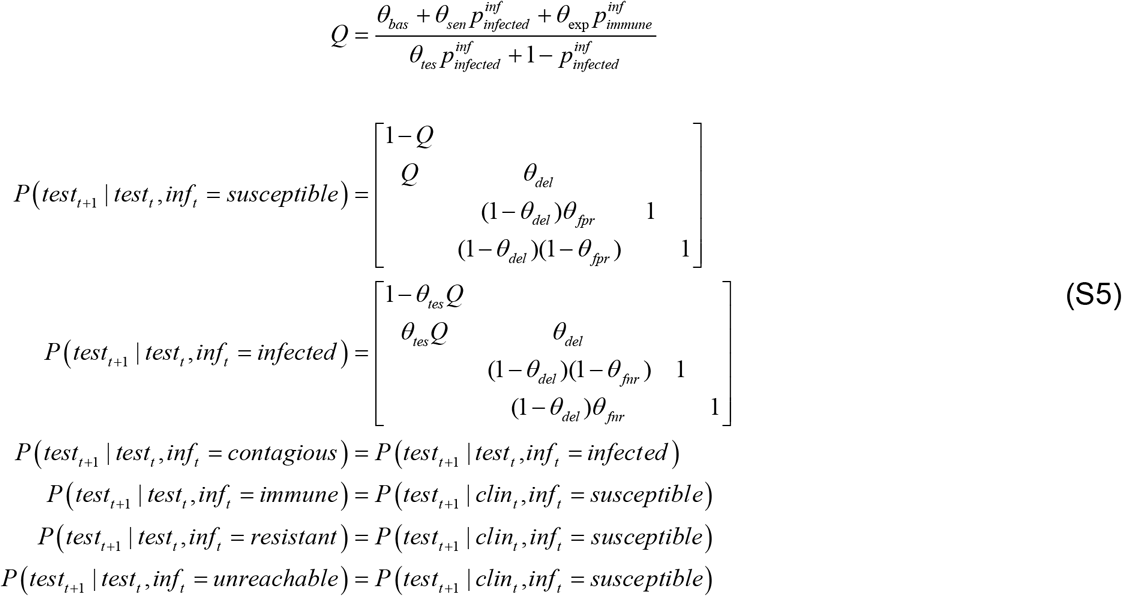

The transition probabilities here are parameterised in terms of test availability (*θ*_*bas*_, *θ*_*sen*_ and *θ*_*exp*_) and the probability that I would have been tested anyway, which is relatively smaller if I am asymptomatic (*θ*_*tes*_). Once tested, I will keep waiting for test results with a delay *θ*_*del*_. After this, I can only go into *positive* or *negative* test states, depending upon whether I have the virus (*i*.*e*., *infected* or *infectious*) or not. Note that specificity and sensitivity of PCR tests are explicitly modelled through false positive/negative rate parameters *θ*_*fpr*_ and *θ*_*fnr*_, respectively (Wang *et al*., 2020).

## Appendix 2

The reliability of our model-based predictions currently depends upon how accurate the fit of the LIST model to region-to-region data time series was. Figure S1 below shows the average percentage of explained variance (across all 4 time series) for each metropolitan French region.

**Figure S1:**
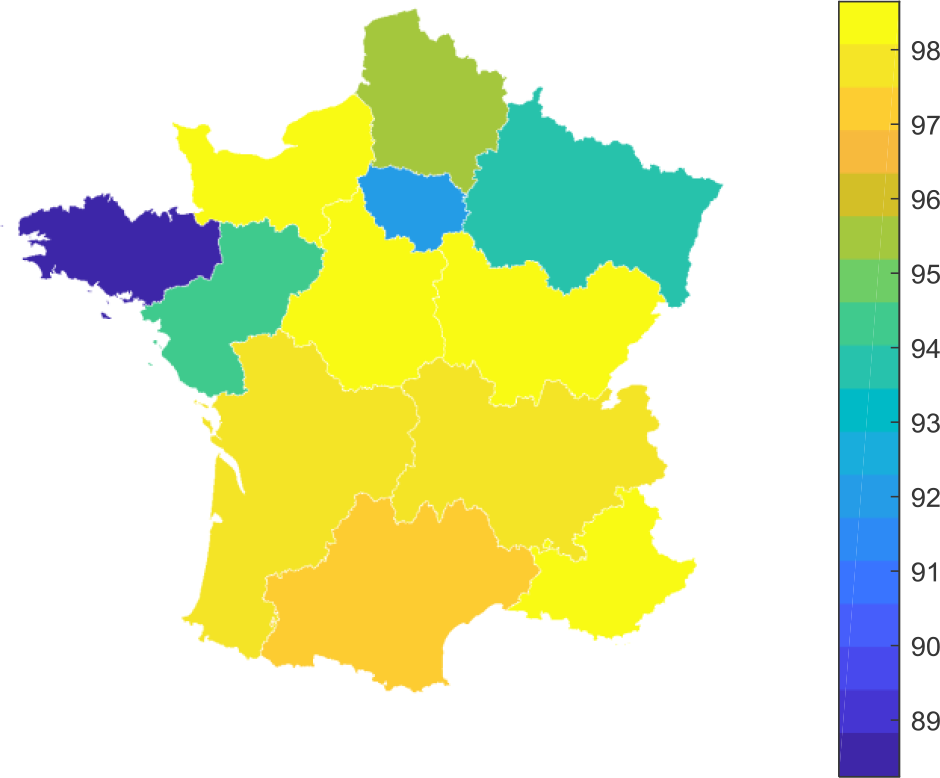
Fit accuracy of the modified LIST model. The average percentage of variance explained (across death rates, positive case rates, remission rates and ICU occupancy) is shown for all metropolitan French regions.

One can see that the LIST model explained more than 95% of the variance in the 4 timeseries for all regions, except *Bretagne* and *Ile-de-France*. The likely reason for the smaller fit accuracy in these regions is idiosyncratic. For *Bretagne*, very few testing laboratories have actually registered in the national reporting network. The consequence of this is that positive cases are under-sampled. For *Ile-de-France*, data are slightly less reliable than in other regions because the region temporarily handled many patients suffering from severe forms of COVID infection, who were shipped from and to other regions in France during the peak of the outbreak.

At this stage, suffice it to say that the model-based predictions and inferences in these two regions may have relatively lower reliability.

## Footnotes

1 “Removed” individuals are either cured (immune) or dead.

2 This is a stable fixed point that corresponds to a successful mitigation strategy, in which everybody has acquired some protective immunity. There also exists an unstable fixed point in which the number of infectious people is zero. This corresponds to a successful suppression strategy (that may have been obtained in places like New Zealand).

3 For example, many mass gathering shave been observed in all major big French cities (https://www.francetvinfo.fr/sante/maladie/coronavirus/une-fete-de-la-musique-sans-trop-de-precautions-pour-oublier-le-coronavirus_4017437.html). Also, the online smartphone app that was meant to regulate quarantining on a voluntary basis has had very limited success until now (https://www.lemonde.fr/pixels/article/2020/06/23/application-stopcovid-14-personnes-averties-en-trois-semaines_6043915_4408996.html).

4 Here, being at “work” essentially means being neither at home, at the hospital or in the morgue (cf., *e*.*g*., children at school).

5 Astute readers will notice a few minor changes from the original model inversion scheme proposed by Friston *et al*. (Friston *et al*., 2020a). In particular, the hard, sigmoid constraint on transition probability parameters ensures that these cannot be greater than one.

6 Among the 1,700 people on board, only 1,046 people were infected (all were tested). See, *e*.*g*., https://www.lesechos.fr/industrie-services/air-defense/coronavirus-lenquete-sur-la-contagion-des-marins-du-porte-avions-charles-de-gaulle-na-releve-aucune-faute-1202063

7 Out of the 3,711 people on board, 700 were infected, 18% of which were asymptomatic. See, e.g., https://www.nature.com/articles/d41586-020-00885-w.

8 In the LIST model, unknown parameters capture the number of ‘at-home’ and ‘at-work’ person-to-person contacts, respectively. Importantly, the former is smaller than the latter. Hence, increasing the proportion of ‘at-work’ people mechanically increase the number of total contacts.

9 This can be done very simply by propagating the adjacency matrix until it reaches a fixed-point.

10 As a reference point, the estimated steady-state marginal probability of being at work in France is about 1/4 (see Figure 12 below). Note that this is rather reasonable, given that (i) not everyone is active, and (ii) people typically work 8 hours a day.

11 World Health Organization (https://www.who.int/).

12 European Centre for Disease Prevention and Control (https://www.ecdc.europa.eu/en).

13 Note that marginal probability 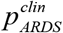 on the third line of Equation 1 is evaluated on the day before, such 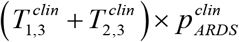 that is the current transition rate towards asymptomatic or symptomatic cases.

14 All these datasets are available from https://www.data.gouv.fr/en/datasets/donnees-hospitalieres-relatives-a-lepidemie-de-covid-19/ and https://www.data.gouv.fr/en/datasets/donnees-relatives-aux-tests-de-depistage-de-covid-19-realises-en-laboratoire-de-ville/. We discarded data from non-metropolitan regions because the exported data were unreliable (high rate of missing data, low density of testing laboratories that report COVID test results, etc).

15 FWHM = Full Width Half Maximum.

16 Estimates of mortality rates are obtained by dividing the cumulated count of deceased people by the corresponding population size. Estimates of prevalence rates are obtained by dividing the cumulated count of positive tests by the cumulated count of total tests. In Figure 13, these estimates are based on available data from Santé Publique France.

17 Current estimates of the false negative rate of COVID (PCR-based) testing technologies could be as high as 30%.

18 In fact, an epidemiological study recently estimated (using an extended SIR model) that about 60,000 lives were saved by the French lockdown (https://www.ehesp.fr/en/2020/04/28/a-study-by-researchers-from-ehesp-french-school-of-public-health-shows-that-lockdown-saved-at-least-60-000-lives-in-france/).

19 This estimate does not account for telecommuting home-workers.

20 https://www.fil.ion.ucl.ac.uk/spm/ and https://mbb-team.github.io/VBA-toolbox/.

## References

Arevalo-Rodriguez, I., Buitrago-Garcia, D., Simancas-Racines, D., Zambrano-Achig, P., Campo, R. del, Ciapponi, A., Sued, O., Martinez-Garcia, L., Rutjes, A., Low, N., et al. (2020). False-negative results of initial RT-PCR assays for COVID-19: a systematic review. MedRxiv 2020.04.16.20066787.

Ashburner, J. (2012). SPM: A history. Neuroimage 62–248, 791–800.

Balabdaoui, F., and Mohr, D. (2020). Age-stratified model of the COVID-19 epidemic to analyze the impact of relaxing lockdown measures: nowcasting and forecasting for Switzerland. MedRxiv 2020.05.08.20095059.

Bao, L., Deng, W., Gao, H., Xiao, C., Liu, J., Xue, J., Lv, Q., Liu, J., Yu, P., Xu, Y., et al. (2020). Reinfection could not occur in SARS-CoV-2 infected rhesus macaques. BioRxiv 2020.03.13.990226.

Bi, Q., Wu, Y., Mei, S., Ye, C., Zou, X., Zhang, Z., Liu, X., Wei, L., Truelove, S.A., Zhang, T., et al. (2020). Epidemiology and transmission of COVID-19 in 391 cases and 1286 of their close contacts in Shenzhen, China: a retrospective cohort study. Lancet Infect. Dis. 0.

Bunyavanich, S., Do, A., and Vicencio, A. (2020). Nasal Gene Expression of Angiotensin-Converting Enzyme 2 in Children and Adults. JAMA 323, 2427–2429.

Daunizeau, J. (2018). The variational Laplace approach to approximate Bayesian inference. ArXiv170302089 Q-Bio Stat.

Daunizeau, J., David, O., and Stephan, K.E. (2011). Dynamic causal modelling: a critical review of the biophysical and statistical foundations. NeuroImage 58, 312–322.

Daunizeau, J., Adam, V., and Rigoux, L. (2014). VBA: A Probabilistic Treatment of Nonlinear Models for Neurobiological and Behavioural Data. PLoS Comput Biol 10, e1003441.

Daunizeau, J., Moran, R.J., Mattout, J., and Friston, K. (2020). On the reliability of model-based predictions in the context of the current COVID epidemic event: impact of outbreak peak phase and data paucity. MedRxiv 2020.04.24.20078485.

Domenico, L.D., Pullano, G., Sabbatini, C.E., Boëlle, P.-Y., and Colizza, V. (2020a). Expected impact of lockdown in Île-de-France and possible exit strategies. MedRxiv 2020.04.13.20063933.

Domenico, L.D., Pullano, G., Sabbatini, C.E., Boëlle, P.-Y., and Colizza, V. (2020b). Expected impact of reopening schools after lockdown on COVID-19 epidemic in Île-de-France. MedRxiv 2020.05.08.20095521.

Edridge, A.W., Kaczorowska, J.M., Hoste, A.C., Bakker, M., Klein, M., Jebbink, M.F., Matser, A., Kinsella, C., Rueda, P., Prins, M., et al. (2020). Coronavirus protective immunity is short-lasting. MedRxiv 2020.05.11.20086439.

Eliasmith, C., and Trujillo, O. (2014). The use and abuse of large-scale brain models. Curr. Opin. Neurobiol. 25, 1–6.

Eubank, S., Eckstrand, I., Lewis, B., Venkatramanan, S., Marathe, M., and Barrett, C.L. (2020). Commentary on Ferguson, et al., “Impact of Non-pharmaceutical Interventions (NPIs) to Reduce COVID-19 Mortality and Healthcare Demand.” Bull. Math. Biol. 82.

Flaxman, S. (2020). Report 13 - Estimating the number of infections and the impact of non-pharmaceutical interventions on COVID-19 in 11 European countries.

Friston, K. (2012). Ten ironic rules for non-statistical reviewers. NeuroImage 61, 1300–1310.

Friston, K., Mattout, J., Trujillo-Barreto, N., Ashburner, J., and Penny, W. (2007). Variational free energy and the Laplace approximation. NeuroImage 34, 220–234.

Friston, K.J., Preller, K.H., Mathys, C., Cagnan, H., Heinzle, J., Razi, A., and Zeidman, P. (2017). Dynamic causal modelling revisited. NeuroImage.

Friston, K.J., Parr, T., Zeidman, P., Razi, A., Flandin, G., Daunizeau, J., Hulme, O.J., Billig, A.J., Litvak, V., Moran, R.J., et al. (2020a). Dynamic causal modelling of COVID-19. 200404463 Q-Bio.

Friston, K.J., Parr, T., Zeidman, P., Razi, A., Flandin, G., Daunizeau, J., Hulme, O.J., Billig, A.J., Litvak, V., Price, C.J., et al. (2020b). Effective immunity and second waves: a dynamic causal modelling study. 200609429 Q-Bio.

Friston, K.J., Parr, T., Zeidman, P., Razi, A., Flandin, G., Daunizeau, J., Hulme, O.J., Billig, A.J., Litvak, V., Price, C.J., et al. (2020c). Second waves, social distancing, and the spread of COVID-19 across America.

Friston, K.J., Parr, T., Zeidman, P., Razi, A., Flandin, G., Daunizeau, J., Hulme, O.J., Billig, A.J., Litvak, V., Price, C.J., et al. (2020d). Tracking and tracing in the UK: a dynamic causal modelling study. 200507994 Q-Bio.

Gandhi, M., Yokoe, D.S., and Havlir, D.V. (2020). Asymptomatic Transmission, the Achilles’ Heel of Current Strategies to Control Covid-19. N. Engl. J. Med. 382, 2158–2160.

Grifoni, A., Weiskopf, D., Ramirez, S.I., Mateus, J., Dan, J.M., Moderbacher, C.R., Rawlings, S.A., Sutherland, A., Premkumar, L., Jadi, R.S., et al. (2020). Targets of T Cell Responses to SARS-CoV-2 Coronavirus in Humans with COVID-19 Disease and Unexposed Individuals. Cell.

Hazem, Y., Natarajan, S., and Berikaa, E. (2020). Hasty Reduction of COVID-19 Lockdown Measures Leads to the Second Wave of Infection. MedRxiv 2020.05.23.20111526.

Inoue, H., and Todo, Y. (2020). The Propagation of the Economic Impact through Supply Chains: The Case of a Mega-City Lockdown against the Spread of COVID-19 (Rochester, NY: Social Science Research Network).

Johnson, N.P.A.S., and Mueller, J. (2002). Updating the accounts: global mortality of the 1918-1920 “Spanish” influenza pandemic. Bull. Hist. Med. 76, 105–115.

Kermack, W.O., McKendrick, A.G., and Walker, G.T. (1927). A contribution to the mathematical theory of epidemics. Proc. R. Soc. Lond. Ser. Contain. Pap. Math. Phys. Character 115, 700–721.

Kissler, S.M., Tedijanto, C., Goldstein, E., Grad, Y.H., and Lipsitch, M. (2020). Projecting the transmission dynamics of SARS-CoV-2 through the postpandemic period. Science.

Lau, H., Khosrawipour, V., Kocbach, P., Mikolajczyk, A., Schubert, J., Bania, J., and Khosrawipour, T. (2020). The positive impact of lockdown in Wuhan on containing the COVID-19 outbreak in China. J. Travel Med. 27.

Lavezzo, E., Franchin, E., Ciavarella, C., Cuomo-Dannenburg, G., Barzon, L., Vecchio, C.D., Rossi, L., Manganelli, R., Loregian, A., Navarin, N., et al. (2020). Suppression of COVID-19 outbreak in the municipality of Vo, Italy. MedRxiv 2020.04.17.20053157.

Lawton, G. (2020). How do we leave lockdown? New Sci. 246, 10–12.

Lebreton, M., Bavard, S., Daunizeau, J., and Palminteri, S. (2019). Assessing inter-individual differences with task-related functional neuroimaging. Nat. Hum. Behav. 3, 897–905.

Leung, K., Wu, J.T., Liu, D., and Leung, G.M. (2020). First-wave COVID-19 transmissibility and severity in China outside Hubei after control measures, and second-wave scenario planning: a modelling impact assessment. The Lancet 395, 1382–1393.

Lin, G., Strauss, A.T., Pinz, M., Martinez, D.A., Tseng, K.K., Schueller, E., Gatalo, O., Yang, Y., Levin, S.A., Klein, E.Y., et al. (2020). Explaining the Bomb-Like Dynamics of COVID-19 with Modeling and the Implications for Policy. MedRxiv 2020.04.05.20054338.

Martini, M., Gazzaniga, V., Bragazzi, N.L., and Barberis, I. (2019). The Spanish Influenza Pandemic: a lesson from history 100 years after 1918. J. Prev. Med. Hyg. 60, E64–E67.

Moghadas, S.M., Shoukat, A., Fitzpatrick, M.C., Wells, C.R., Sah, P., Pandey, A., Sachs, J.D., Wang, Z., Meyers, L.A., Singer, B.H., et al. (2020). Projecting hospital utilization during the COVID-19 outbreaks in the United States. Proc. Natl. Acad. Sci.

Moxnes, J.F., and Christophersen, O.A. (2008). The Spanish flu as a worst case scenario? Microb. Ecol. Health Dis. 20, 1–26.

Ng, K., Faulkner, N., Cornish, G., Rosa, A., Earl, C., Wrobel, A., Benton, D., Roustan, C., Bolland, W., Thompson, R., et al. (2020). Pre-existing and de novo humoral immunity to SARS-CoV-2 in humans. BioRxiv 2020.05.14.095414.

Palminteri, S., Wyart, V., and Koechlin, E. (2017). The Importance of Falsification in Computational Cognitive Modeling. Trends Cogn. Sci. 21, 425–433.

Pastor-Satorras, R., and Vespignani, A. (2001). Epidemic Spreading in Scale-Free Networks. Phys. Rev. Lett. 86, 3200–3203.

Peto, J., Alwan, N.A., Godfrey, K.M., Burgess, R.A., Hunter, D.J., Riboli, E., Romer, P., Buchan, I., Colbourn, T., Costelloe, C., et al. (2020). Universal weekly testing as the UK COVID-19 lockdown exit strategy. The Lancet 395, 1420–1421.

Phillips, H. (2014). The Recent Wave of ‘Spanish’ Flu Historiography. Soc. Hist. Med. 27, 789–808.

Prompetchara, E., Ketloy, C., and Palaga, T. (2020). Immune responses in COVID-19 and potential vaccines: Lessons learned from SARS and MERS epidemic. Asian Pac. J. Allergy Immunol. 38, 1–9.

Reid, A.H., Taubenberger, J.K., and Fanning, T.G. (2001). The 1918 Spanish influenza:integrating history and biology. Microbes Infect. 3, 81–87.

Roux, J., Massonnaud, C., and Crépey, P. (2020). COVID-19: One-month impact of the French lockdown on the epidemic burden. MedRxiv 2020.04.22.20075705.

Salje, H., Kiem, C.T., Lefrancq, N., Courtejoie, N., Bosetti, P., Paireau, J., Andronico, A., Hozé, N., Richet, J., Dubost, C.-L., et al. (2020). Estimating the burden of SARS-CoV-2 in France. Science.

Sarma, U., and Ghosh, B. (2020). Quantitative modeling and analysis show country-specific optimization of quarantine measures can potentially circumvent COVID19 infection spread post lockdown. MedRxiv 2020.05.20.20107169.

Small, M., Tse, C.K., and Walker, D.M. (2006). Super-spreaders and the rate of transmission of the SARS virus. Phys. Nonlinear Phenom. 215, 146–158.

Spinney, L. (2018). Pale rider: the Spanish flu of 1918 and how it changed the world (London: Vintage).

Thornton, J. (2020). Covid-19: A&E visits in England fall by 25% in week after lockdown. BMJ 369.

Wang, W., Xu, Y., Gao, R., Lu, R., Han, K., Wu, G., and Tan, W. (2020). Detection of SARS-CoV-2 in Different Types of Clinical Specimens. JAMA 323, 1843–1844.

Watson, J., Whiting, P.F., and Brush, J.E. (2020). Interpreting a covid-19 test result. BMJ 369.

Xiao Fan Wang, and Guanrong Chen (2003). Complex networks: small-world, scale-free and beyond. IEEE Circuits Syst. Mag. 3, 6–20.

Xu, S., and Li, Y. (2020). Beware of the second wave of COVID-19. The Lancet 395, 1321–1322.

Zheng, M., Gao, Y., Wang, G., Song, G., Liu, S., Sun, D., Xu, Y., and Tian, Z. (2020). Functional exhaustion of antiviral lymphocytes in COVID-19 patients. Cell. Mol. Immunol. 17, 533–535.

